# Genetic association studies using disease liabilities from deep neural networks

**DOI:** 10.1101/2023.01.18.23284383

**Authors:** Lu Yang, Marie C. Sadler, Russ B. Altman

## Abstract

The case-control study is a widely used method for investigating the genetic underpinnings of binary traits. However, long-term, prospective cohort studies often grapple with absent or evolving health-related outcomes. Here, we propose two methods, *liability* and *meta*, for conducting genome-wide association study (GWAS) that leverage disease liabilities calculated from deep patient phenotyping. Analyzing 38 common traits in ∼300,000 UK Biobank participants, we identified an increased number of loci compared to the conventional case-control approach, with high replication rates in larger external GWAS. Further analyses confirmed the disease-specificity of the genetic architecture with the meta method demonstrating higher robustness when phenotypes were imputed with low accuracy. Additionally, polygenic risk scores based on disease liabilities more effectively predicted newly diagnosed cases in the 2022 dataset, which were controls in the earlier 2019 dataset. Our findings demonstrate that integrating high-dimensional phenotypic data into deep neural networks enhances genetic association studies while capturing disease-relevant genetic architecture.

## Introduction

The advent of biobanks has greatly reduced the cost and difficulty of collecting large genome-wide data^1,2^, and has enabled a more comprehensive understanding of the genetic basis of human health and disease^3^. The UK Biobank (UKBB) database comprises genetic data for about 500,000 participants together with socio-demographic, lifestyle and health-related phenotypes^4^. Genome-wide association studies (GWAS) conducted on biobanks such as the UKBB have provided new insights into the genetic architecture of a wide range of quantitative traits and complex diseases^5^. However, while these studies have identified many disease susceptibility loci, the proportion of phenotypic variance that is captured remains limited^6^.

To unravel the role of genetics in disease, studies typically compare patients with the disease (*cases*) to healthy individuals (*controls*). However, patient cohorts may progress beyond a dichotomous setting of “diseased”and “non-diseased”. Healthy patients can become diseased years down the line and GWAS studies explicitly modelling age-at-onset showed an increase in statistical power compared to case-control (CC) studies^7–9^, although the difference becomes small for low-prevalence diseases^10,11^. Time-to-event analyses are computationally expensive and alternative approaches have focused on integrating family history to capture the underlying disease liability. For instance, GWAS by proxy (GWAX) where individuals who reported cases within family members are included as cases themselves have reported increased numbers of new risk loci^12^. GWAX has been particularly useful in the study of Alzheimer’s disease where 46,828 proxy-cases in addition to 2,447 diagnosed cases in the UKBB contributed to increased statistical power^13,14^. Going beyond a binary phenotype, more refined models have integrated participant’s family history to derive a continuous genetic liability phenotype where CC and family history configurations are differentiated more appropriately^15^.

While a participant’s age may mask a future disease onset, disease diagnoses in biobanks can also be missing. To circumvent the issue of mislabeling cases as controls, methods have emerged to explore the joint contribution of family history, clinical measures, and lifestyle factors to model disease liabilities. Machine learning (ML) techniques have advanced in recent years, revealing patterns in massive, high-dimensional datasets capturing non-linear relationships^16,17^. Several ML models that predict disease probabilities based on available biobank-scale features have demonstrated increased statistical power in GWAS while remaining disease-specific^18–21^. More recently, a deep neural network was employed for phenotype imputation which had increased phenotype imputation accuracy compared to standard ML models such as K-Nearest Neighbors and random forests, and could accommodate thousands of traits available in the UKBB^22^.

In this study, we aimed to enhance the power of GWAS and improve genetic predictions by estimating disease liabilities through a deep learning framework called POPDx^23^. POPDx computes joint semantic and structure-based embeddings of phenotypes in the UKBB, utilizing the Human Disease Ontology. It calculates phenotype liability scores for over 1,500 traits in the UK Biobank, encompassing 1,538 phecodes and 12,803 ICD-10 codes. Based on these disease liability scores we computed GWAS following two different approaches, one employing liability scores across all individuals (*liability*) and another using scores for controls only while incorporating case-control status (*meta*). Applying this framework to 38 traits, we not only increased the GWAS power compared to traditional case-control approaches but also observed higher polygenic risk scores (PRS) for newly diagnosed cases. As disease liability scores borrow information from related phenotypes, genetic effects non-specific to the disease under investigation may be identified. However, through genetic correlations and external GWAS comparisons, we demonstrate that the genetic architecture derived from the liability methods preserved disease specificity. Overall, our findings highlight that conditions with frequent missing diagnoses particularly benefit from phenotype imputations via deep learning.

## Results

### Overview of the analysis

Following the extraction of multiple layers of information, including laboratory tests, physical measurements, answered questionnaires and interviews about the socio-demographic status, lifestyle, and family history, we computed disease liability scores for 261,759 participants of the UKBB. This step was performed with training a deep learning framework (POPDx)^23^ (Figure 1A; Figure S1). Across the 38 traits, POPDx demonstrated very high accuracies with area under the receiver operating characteristic (AUROC) values of at least 75% and high area under the precision-recall curve (AUPRC) as shown in Table 1 (sensitivity and specificities at different thresholds are shown in Tables S3-4). Next, these trait predictions were employed as disease liability scores for downstream genetic analysis, which were then compared with the traditional CC approach. Thus, we computed GWAS i) in an ordinary CC logistic regression framework (*binary*), ii) by treating phenotype predictions as a quantitative outcome trait (*liability*), and iii) by meta-analyzing the binary GWAS and a quantitative trait (phenotype predictions) GWAS on controls only (*meta*) capitalizing on the independence of these two analysis sets (Figure 1B). The utilization of the control individuals in both the quantitative control and binary GWAS could suggest sample overlap which would inflate genetic effect estimates. However, the null phenotypic correlation between the overlapping control samples ensures an effective sample overlap of zero and thus no genetic inflation which we further demonstrate in a simulation analysis (Supplementary Note). We then compared disease-associated risk loci identified by the liability approaches to the associations from the CC method. As large association counts could arise from both disease-specific and nonspecific components, we further conducted multiple analyses to determine disease specificity. First, we verified whether liability- and meta-only loci were significant in external GWAS. Second, we computed genetic correlations between liability, meta and binary GWAS while also investigating potential sources of nonspecific genetic effects. Lastly, we estimated PRS effects using the liability scale as well as the original CC status (Figure 1C). We then focused on new cases that have emerged in a more recent release of the UKBB and compared their PRS percentiles obtained from the liability and CC phenotypes. Overall, this study aimed at quantifying the extent to which deep learning-based phenotyping contributes to genetic prediction by discovering new disease variants and enhancing PRS derivations, all while preserving disease-specificity.

**Figure 1.**
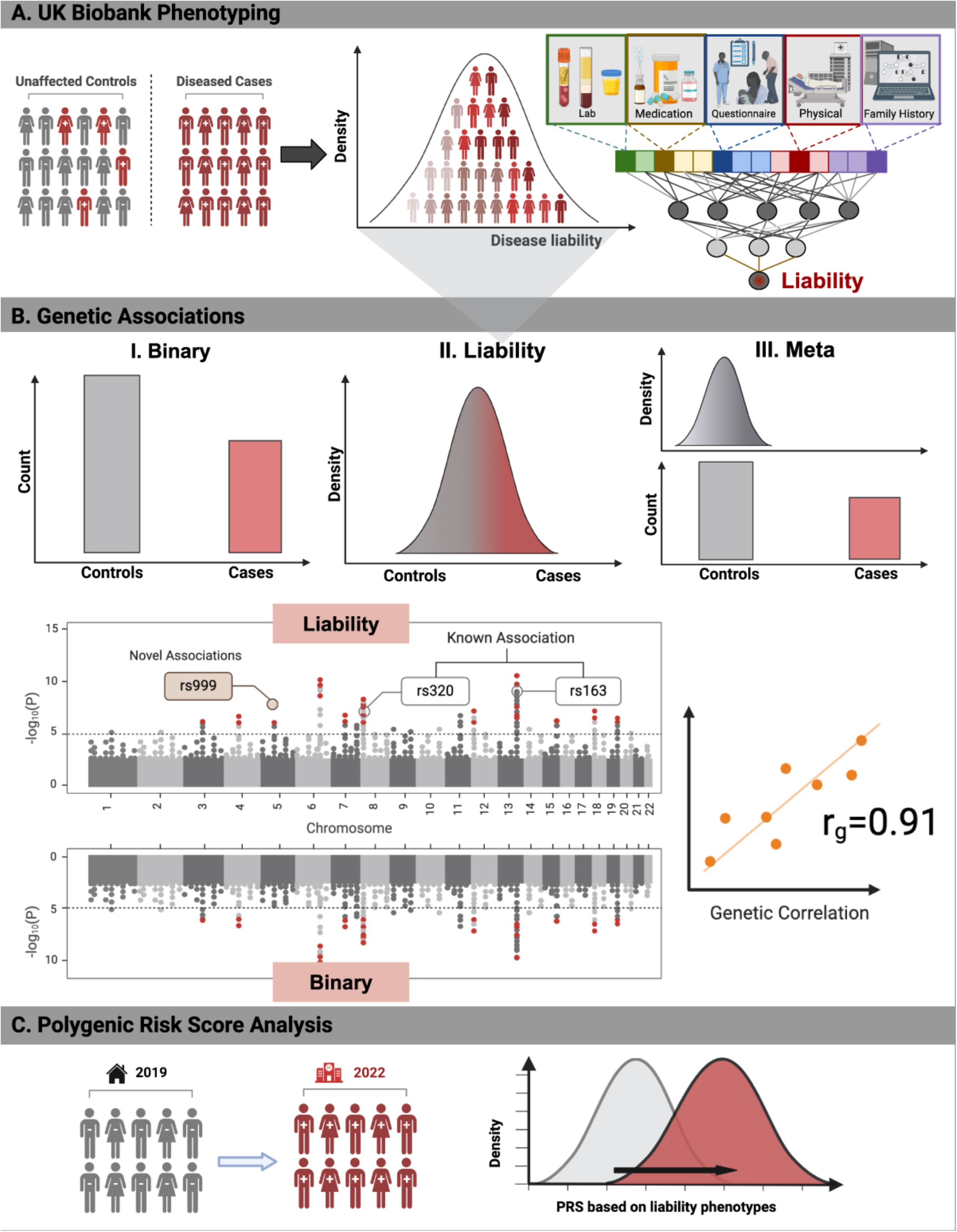
Overview of the analysis. (A) Patient phenotyping in UK Biobank is the first step in this study’s workflow. After extracting the data, we preprocess available patient information such as laboratory results, medication intake, answered questionnaires, medical records, and family history. A deep neural network (POPDx) is then trained on the one-hot encoding matrix of patient embeddings with the corresponding binary (case-control) diagnostic labels to obtain disease liabilities for each disease of interest. (B) Three different methods are used to conduct genome-wide association studies (GWAS) of 38 complex traits, including (i) case and control (CC) logistic regression GWAS (*binary*), (ii) quantitative GWAS on liability phenotype scores (*liability*), and (iii) meta-analysis of a quantitative liability GWAS on controls and CC GWAS on all individuals (*meta*). To test disease-specificities of the liability methods, we compare GWAS signals between methods and validated novel associations (i.e., associations missed by the binary method) in external GWAS. Furthermore, we conduct genetic correlations between liability, binary and external GWAS. (C) We identify newly diagnosed cases in the 2022 compared to the 2019 release of ICD-10 codes and compare the polygenic risk scores of these newly identified cases calculated from the liability and CC status of the patients. Figure created with BioRender.com.

**Table 1.** Performance of the deep learning model POPDx on 38 traits in the UK Biobank (UKBB). AUROC, area under the receiver operating characteristic; AUPRC, area under precision recall curve; Prevalence, disease prevalence in the UKBB.

| Phenotype | Phecode | AUROC | AUPRC | Prevalence |
| --- | --- | --- | --- | --- |
| Essential hypertension | 401.1 | 0.9409 | 0.8659 | 0.2419 |
| Transient cerebral ischemia | 433.31 | 0.8935 | 0.7717 | 0.227 |
| Abdominal pain | 785 | 0.8042 | 0.4218 | 0.1252 |
| Hypercholesterolemia | 272.11 | 0.9156 | 0.6309 | 0.1037 |
| Diverticulosis | 562.1 | 0.9248 | 0.6412 | 0.0891 |
| Diaphragmatic hernia | 550.2 | 0.8563 | 0.4905 | 0.0843 |
| Asthma | 495 | 0.9511 | 0.7584 | 0.0825 |
| Disorders of iris and ciliary body | 379.5 | 0.9563 | 0.7866 | 0.0786 |
| Hyperplasia of prostate | 600 | 0.9682 | 0.6373 | 0.078 |
| Hemorrhoids | 455 | 0.9299 | 0.6525 | 0.0735 |
| Benign neoplasm of colon | 208 | 0.9517 | 0.7487 | 0.0666 |
| Cancer of brain | 191.11 | 0.9332 | 0.7008 | 0.063 |
| Coronary atherosclerosis | 411.4 | 0.9836 | 0.8995 | 0.0613 |
| Type 2 diabetes | 250.2 | 0.9511 | 0.7527 | 0.0603 |
| Inguinal hernia | 550.1 | 0.9519 | 0.7196 | 0.06 |
| Malignant neoplasm of female breast | 174.11 | 0.9973 | 0.9601 | 0.0573 |
| Gastritis and duodenitis | 535 | 0.8377 | 0.2928 | 0.0526 |
| Cataract | 366 | 0.9351 | 0.6709 | 0.0515 |
| Allergy/adverse effect of penicillin | 960.2 | 0.7584 | 0.1882 | 0.0507 |
| Circulatory disease NEC | 459.9 | 0.9391 | 0.6251 | 0.0506 |
| Hematuria | 593 | 0.9761 | 0.7691 | 0.0505 |
| Angina pectoris | 411.3 | 0.9682 | 0.7775 | 0.05 |
| Atrial fibrillation and flutter | 427.2 | 0.9348 | 0.6368 | 0.0465 |
| Hypothyroidism NOS | 244.4 | 0.9745 | 0.8332 | 0.0441 |
| Uveitis, noninfectious or NOS | 371.1 | 0.9132 | 0.4644 | 0.0397 |
| Urinary tract infection | 591 | 0.8824 | 0.3409 | 0.0384 |
| Myocardial infarction | 411.2 | 0.9995 | 0.996 | 0.0362 |
| Tobacco use disorder | 318 | 0.9608 | 0.6237 | 0.0341 |
| Obesity | 278.1 | 0.9439 | 0.53 | 0.0337 |
| Reflux esophagitis | 530.14 | 0.8591 | 0.2616 | 0.033 |
| Varicose veins of lower extremity | 454.1 | 0.9909 | 0.9463 | 0.0325 |
| Enthesopathy | 726.1 | 0.9217 | 0.5248 | 0.0319 |
| Ulceration of intestine | 556.1 | 0.9603 | 0.6757 | 0.0307 |
| Osteoarthritis; localized | 740.1 | 0.9133 | 0.4682 | 0.0291 |
| Cholelithiasis | 574.1 | 0.9699 | 0.6862 | 0.0274 |
| Benign neoplasm of skin | 216 | 0.933 | 0.4624 | 0.0271 |
| Alcoholism | 317.1 | 0.8714 | 0.2678 | 0.0267 |
| Peripheral enthesopathies and allied syndromes | 726 | 0.9719 | 0.793 | 0.0265 |

### Liability phenotypes have increased GWAS power

For the 38 traits, we conducted GWAS for the binary, liability and meta methods on ∼600,000 common directly genotyped SNPs. In Figure 2, Manhattan plots for type 2 diabetes (Figure 2A) and asthma (Figure 2B) show the comparison between the liability and binary methods, with novel risk loci being annotated. For type 2 diabetes, 71 independent loci were identified in the liability method of which 9 were in common with the binary method and 21 loci were only found in the binary method. For example, rs7756992 (*CDKAL1*), rs2307111 (*POC5*), rs3810291 (*ZC3H4*), and rs3923113 (*GRB14*) were not identified using the binary method but showed significant associations in liability GWAS, consistent with findings across the literature^24–31^. Likewise, for asthma, 81 independent loci were identified in the liability method of which 19 were in common with the binary method and 8 loci were only found in the binary method. The loci rs1059513 (*STAT6*), rs11071559 (*RORA*), rs61839660 (*IL2RA*), and rs9290877 (*LPP*) were not detected using the binary method but demonstrated significant associations in the liability GWAS, which aligns with findings reported in the literature^32–38^. Across all traits, the liability methods discovered a larger number of independent risk loci than the binary method with 433, 927 and 782 total number of signals identified by the binary, liability, and meta methods, respectively (Figure 3). When comparing the signals unique to the binary and liability methods by incorporating linkage disequilibrium (LD) information of neighboring SNPs (see Methods), we found that 242 of the loci were identified by only the binary method, while 735 and 541 loci significant in the liability and meta approaches, respectively, did not reach genome-wide significance in the binary method (Figure 3).

**Figure 2.**
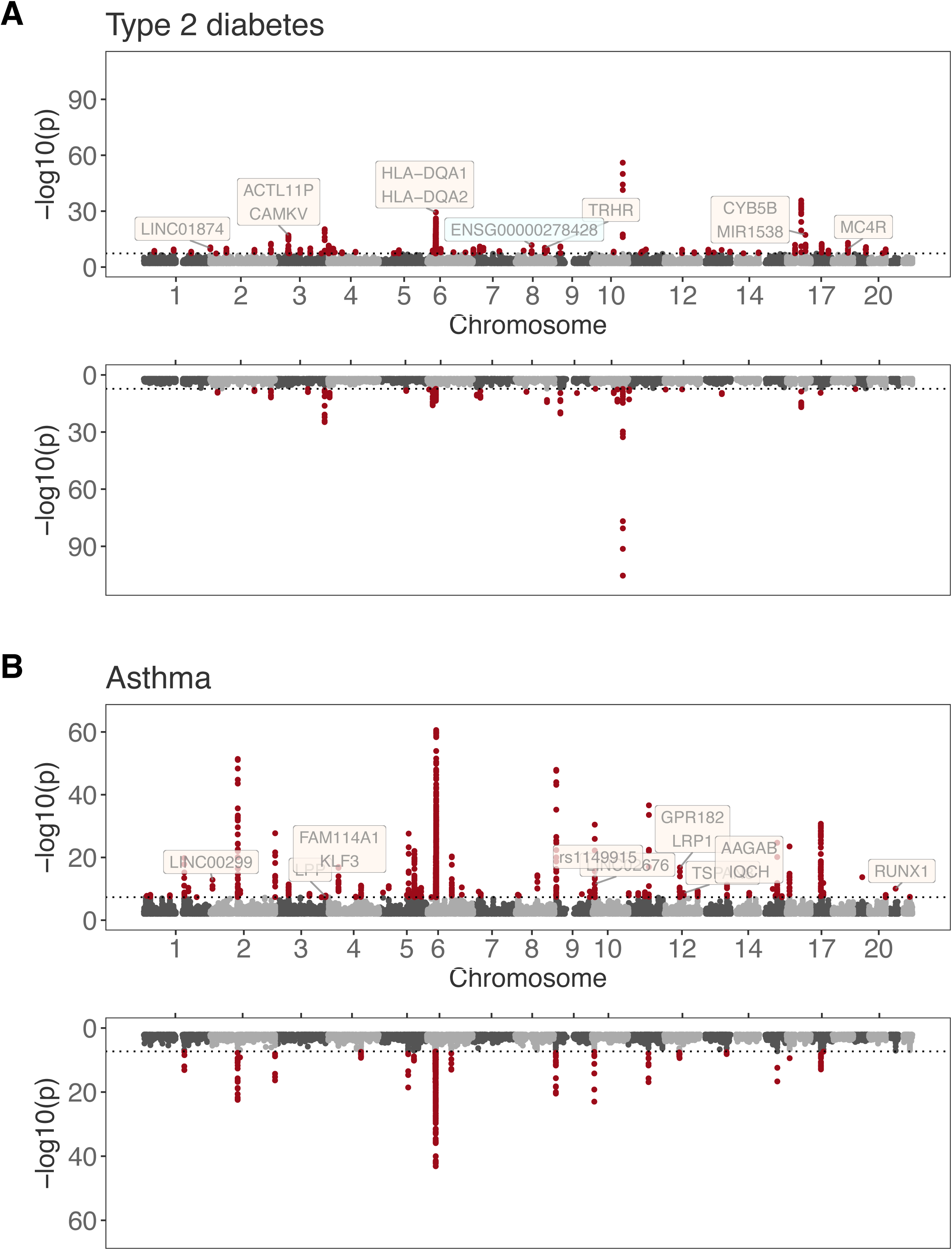
Comparison of GWAS results of type 2 diabetes and asthma between the liability and binary method. Mirrored Manhattan plots of GWAS results for (A) type 2 diabetes and (B) asthma. The liability GWAS results are shown in the top panel and binary results in the bottom panel with loci reaching genome-wide significance plotted in red. Risk loci that did not reach genome-wide significance in the binary method are annotated with the closest gene and highlighted in orange if significant in the external GWAS and blue if not. Chromosome positions are plotted in genomic order on the x-axis and the negative logarithm of p-values (i.e., association strength) on the y-axis. The dotted horizontal line denotes the genome-wide significance threshold of 5 × 10^−8^.

**Figure 3.**
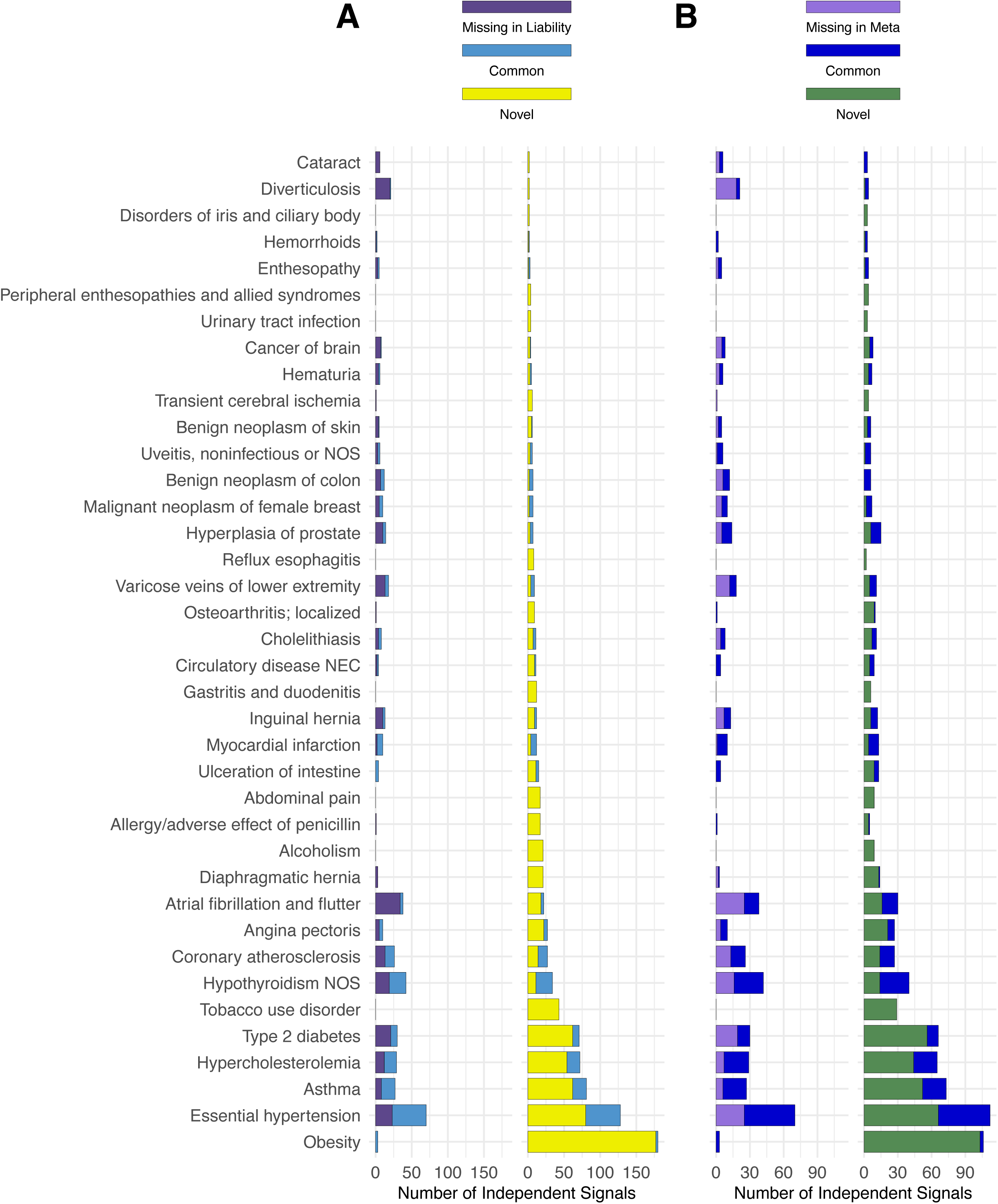
Comparison of the number of independent GWAS loci for the binary, liability and meta methods across the 38 complex traits. (A) Comparison of GWAS signals identified by the binary and liability methods, where ‘Common’ signals reached genome-wide significance in both analyses, ‘Missing in Liability’ signals reached genome-wide significance only in the binary method, and ‘Novel’ signals reached genome-wide significance only in the liability method. (B) Comparison of GWAS signals identified by the binary and meta methods, where ‘Common’ signals reached genome-wide significance in both analyses, ‘Missing in Meta’ signals reached genome-wide significance only in the binary method, and ‘Novel’ signals reached genome-wide significance only in the meta method.

We also calculated SNP-based heritability (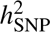) estimates using the BLD-LDAK model which assumes that the expected 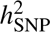 depends on LD, minor allele frequency (MAF) and 66 functional annotations^39^. On the observed scale, 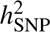 estimates were higher for the liability methods than for the binary method (paired two-sided t-test: p-values of 8.23e-08 and 9.15e-06 for liability and meta, respectively, Table S1). As the observed scale does not reflect the underlying continuous liability of binary traits, we further calculated liability-scale heritability (LSH) estimates for the binary method assuming equal biobank sample as population disease prevalence^40^. After the transformation to the liability scale, heritability estimates of the binary method increased substantially (average 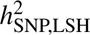 of 0.131 vs. 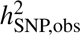 of 0.0328; Table S1) and were higher than for the liability method (average 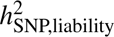 of 0.0974; paired two-sided t-test p-value of 0.0294). However, as estimates of LSH are prone to be inflated and have large estimator variance, especially for diseases with low prevalence^40^, it remains uncertain whether the variance explained by genetics is higher for the binary compared to the liability and meta methods. Nonetheless, compared to the LSH binary heritability estimates, we found that 10 traits—including hypercholesterolemia, obesity, alcoholism, tobacco use disorder, circulatory disease NEC, reflux esophagitis, gastritis and duodenitis, diaphragmatic hernia, urinary tract infection, abdominal pain, and allergy/adverse effect of penicillin—had higher heritability estimates with the liability method (Table S1).

### Novel loci demonstrate high validation rates in external GWAS

We further investigated whether loci identified only in the liability and meta GWAS were trait-specific by verifying their significance level in external GWAS. Since disease liability scores aggregate phenotypes across the whole biobank, the boost in genetic power may come from genetic signals that are associated with related traits but that are non-specific to the studied disease. For this purpose, we consulted GWAS with larger sample sizes, mostly stemming from large meta-analyses, as well as GWAS conducted on quantitative phenotypes matching disease phenotypes (e.g. LDL cholesterol matching hypercholesterolemia and systolic blood pressure matching hypertension; Table S2). Among the eleven traits considered in this analysis, 486 independent SNPs reached genome-wide significance in the liability, but not in the binary GWAS and of these, 432 were available in the external GWAS (Table S20). In total, 330 (76%) SNPs were significant in the larger disease or quantitative trait meta-GWAS at a Bonferroni-corrected p-value threshold of 0.05/432 = 1.16e-4 and 389 (90%) at a nominally significant p-value (256 (59%) SNPs reached genome-wide significance). We repeated the analysis omitting obesity, as under-reported diagnoses resulted in few genome-wide significant hits for this trait in the binary GWAS. Of the remaining 269 liability-specific SNPs, 180 (67%) SNPs were significant in the external GWAS at a Bonferroni-corrected p-value threshold of 0.05/269 = 1.86e-4 and 226 (84%) at a nominally significant p-value (118 (44%) SNPs reached genome-wide significance). Repeating the analysis with the meta method, we found that among the eleven traits, 366 independent SNPs reached genome-wide significance in the meta analysis, but not in the binary GWAS of which 321 were available in the external GWAS (Table S21). In total, 254 (79%) SNPs were significant in the larger disease or quantitative trait meta-GWAS at a Bonferroni-corrected p-value threshold of 0.05/321 = 1.56e-4 and 296 (92%) at a nominally significant p-value (197 (61%) SNPs reached genome-wide significance). Omitting obesity, 167 (73%) of the remaining 229 SNPs were significant in the external GWAS at a Bonferroni-corrected p-value threshold of 0.05/229 = 2.18e-4 and 204 (89%) at a nominally significant p-value (116 (51%) SNPs reached genome-wide significance). Overall, these analyses confirmed that the large majority of loci identified solely by the liability methods were indeed trait-specific.

### Disease liability GWAS remain trait-specific

In addition to validating whether new GWAS hits from liability phenotypes are trait-specific, we calculated genetic correlations with the binary GWAS as well as with external GWAS to estimate whether the polygenic architecture is preserved. In the comparison with the binary GWAS, which are expected to be disease-specific, we observed high genetic correlations across traits (Figure 4A; Table S7). We found lower genetic correlations with the binary GWAS for the liability GWAS of controls only (first step of the meta method, average *r_g_* = 0.646). However, correlations are very high for the liability (average *r_g_* = 0.847) and meta (average *r_g_* = 0.894) GWAS. Next, we tested whether different sets of features could introduce non-specificity. We estimated liability phenotypes by 1) removing all lifestyle-related features (see Method section: Modelling phenotype liabilities) and 2) including only lifestyle-related features. After performing GWAS on these alternative liability phenotypes, we re-estimated genetic correlations. Liability phenotypes derived solely from lifestyle features had much lower genetic correlations with the binary GWAS (average *r_g_* of 0.382, 0.467 and 0.755 for the control liability, liability and meta methods, respectively). This indicates that lifestyle features alone are insufficient for obtaining robust and accurate liability phenotype scores (AUROCs and AUPRCs in Tables S5-6). On the other hand, we found no evidence that lifestyle features reduce trait-specificities. Except for the controls liability GWAS, genetic correlations with the full feature set (average *r_g_*of 0.646, 0.847, and 0.894 for the control liability, liability, and meta methods, respectively) were as high or higher than those without lifestyle features (average *r_g_* of 0.647, 0.842, and 0.894, respectively, when lifestyle features were removed).

**Figure 4.**
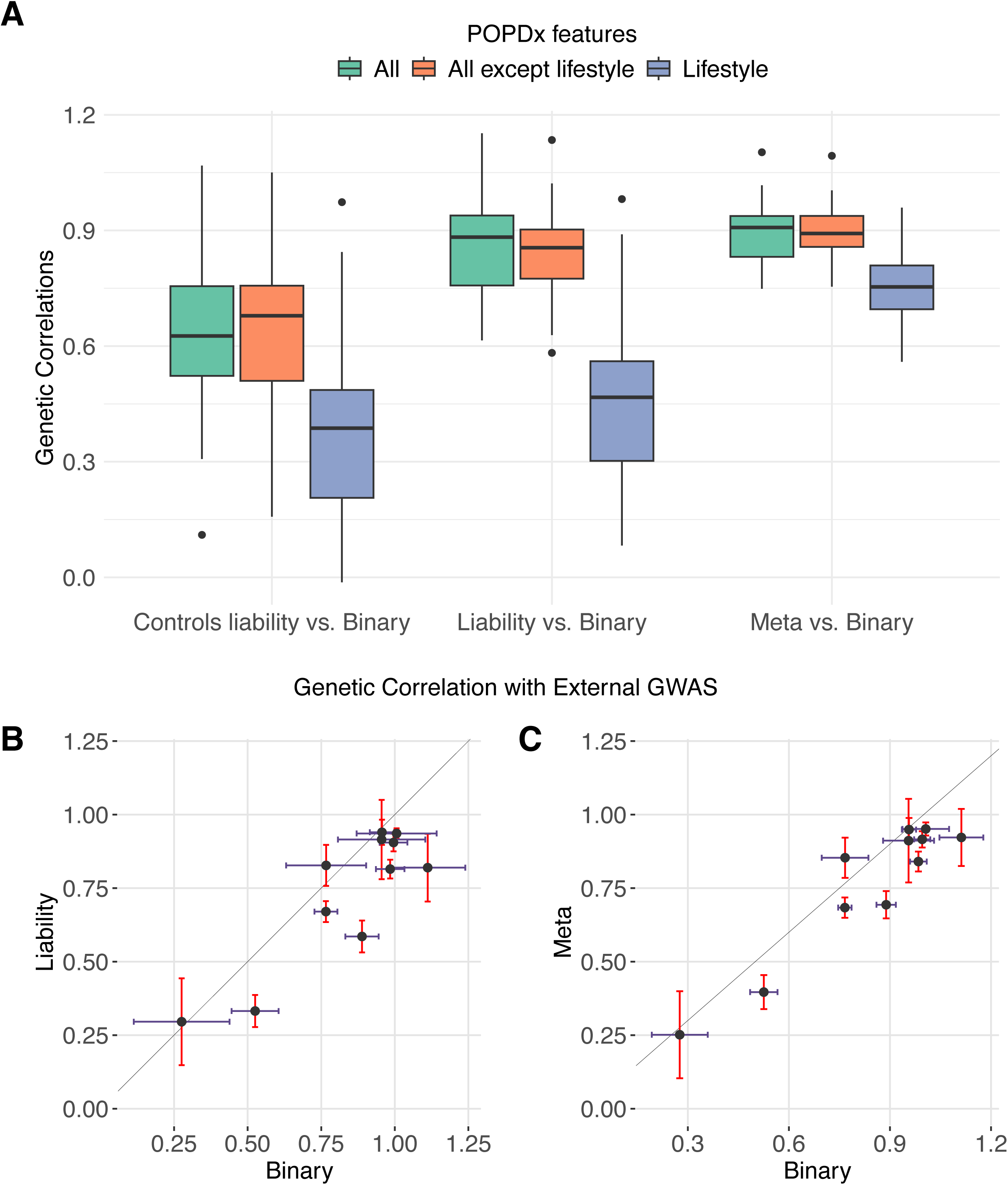
Genetic correlations of liability GWAS with binary and external GWAS. (A) Genetic correlations with binary GWAS across the 38 traits using different POPDx feature sets to calculate liability phenotype scores. The “Quantitative” method refers to GWAS calculated with liability phenotype scores on controls only (first step of the meta method). The boxplots bound the 25th, 50th (median, center), and 75th quantiles. Whiskers range from minima (Q1 - (1.5 x IQR)) to maxima (Q3 + (1.5 x IQR)) with points outside representing potential outliers. Panel (B) and (C) show genetic correlations with external GWAS for the liability (B) and meta (C) methods across 11 traits. Dots represent genetic correlation estimates and error bars the 95% confidence intervals.

Furthermore, we calculated genetic correlations for the binary, liability and meta methods with the external GWAS that were used in the novel SNP validation analysis (Table S2). Genetic correlations were very high (*r_g_>* 0.6) and concordant across the binary, liability and meta methods (Figure 4B-C; Table S8), except for stroke and LDL-cholesterol. However, genetic correlations with external GWAS were higher for the binary (average *r_g_*= 0.839) than for the liability (average *r_g_* = 0.731) and meta (average *r_g_* = 0.761) methods. This higher correlation for the binary method might reflect a bias towards the standard case-control approach, as 9 out of 11 external GWAS included the UKBB as a participating cohort. Despite this, the liability and meta methods demonstrated high robustness and consistency across traits evidencing that the polygenic signals captured by the phenotype imputation methods are relevant to the disease under study.

### Liability derived PRS improve the identification of emerging cases

Placing individuals on a continuous spectrum instead of assigning them a binary CC value holds the promise of capturing who is most at risk for developing the disease. To test this hypothesis, we identified the newly emerged cases in the 2022 release of UKBB diagnosis records compared to the previous release (2019; Figure S2). First, we assessed whether liability scores were indeed higher for emerging cases than for controls which was the case for all traits (Figure S3, Table S9). We next tested whether the same holds for PRS and estimated predictor (i.e., SNP) effects for the binary, liability and meta methods and calculated corresponding PRS. Thus, each individual was assigned a binary, liability and meta PRS. Figure 5 shows how persistent controls (no disease diagnosis in 2019 and 2022), emerging cases (new disease diagnosis in 2022) and cases already defined as such in 2019 rank on the binary and liability PRS scale for six traits (T2D, obesity, hypercholesterolemia, hypertension, asthma and osteoarthritis). For all traits, PRS calculated with liability disease phenotypes, resulted in a more distinct separation of these three patient groups which was the most pronounced for obesity (median liability PRS of −0.0111, 0.197 and 0.510 for persistent controls, emerging cases, and cases, respectively, compared to −0.00727, −0.00303, and 0.203 for binary PRS) followed by T2D (median liability PRS of −0.0207, 0.0818 and 0.399 for persistent controls, emerging cases and cases, respectively, compared to −0.0140, 0.0118 and 0.250 for binary PRS). Across all traits, the average median liability PRS were −0.00789, 0.0280 and 0.174 compared to −0.00904, −0.00158, and 0.130 for binary PRS and −0.00821, 0.0132 and 0.163 for meta PRS for persistent controls, emerging cases and cases, respectively (Figure S5; Tables S10-12). Evaluating the statistical significance of these PRS ranks, we found that emerging cases significantly ranked higher compared to persistent controls in 37/38 traits for liability-derived PRS (p-value *<* 0.05/38 in Wilcoxon rank tests; Table S13) compared to 30/38 traits for binary-derived PRS (Figure 6A; Table S13) with similar results found for meta-derived PRS (37/38 traits; Figure S4; Table S13). When translating the improvement in genetic prediction to PRS percentiles, we found that across diseases emerging cases had on average a 3.20% and 2.25% percentile increase for the liability and meta-derived PRS, respectively, compared to the binary approach (Figure 6B, Tables S14-15) and this difference was significant in 34 and 31 traits, respectively (Table S16).

**Figure 5.**
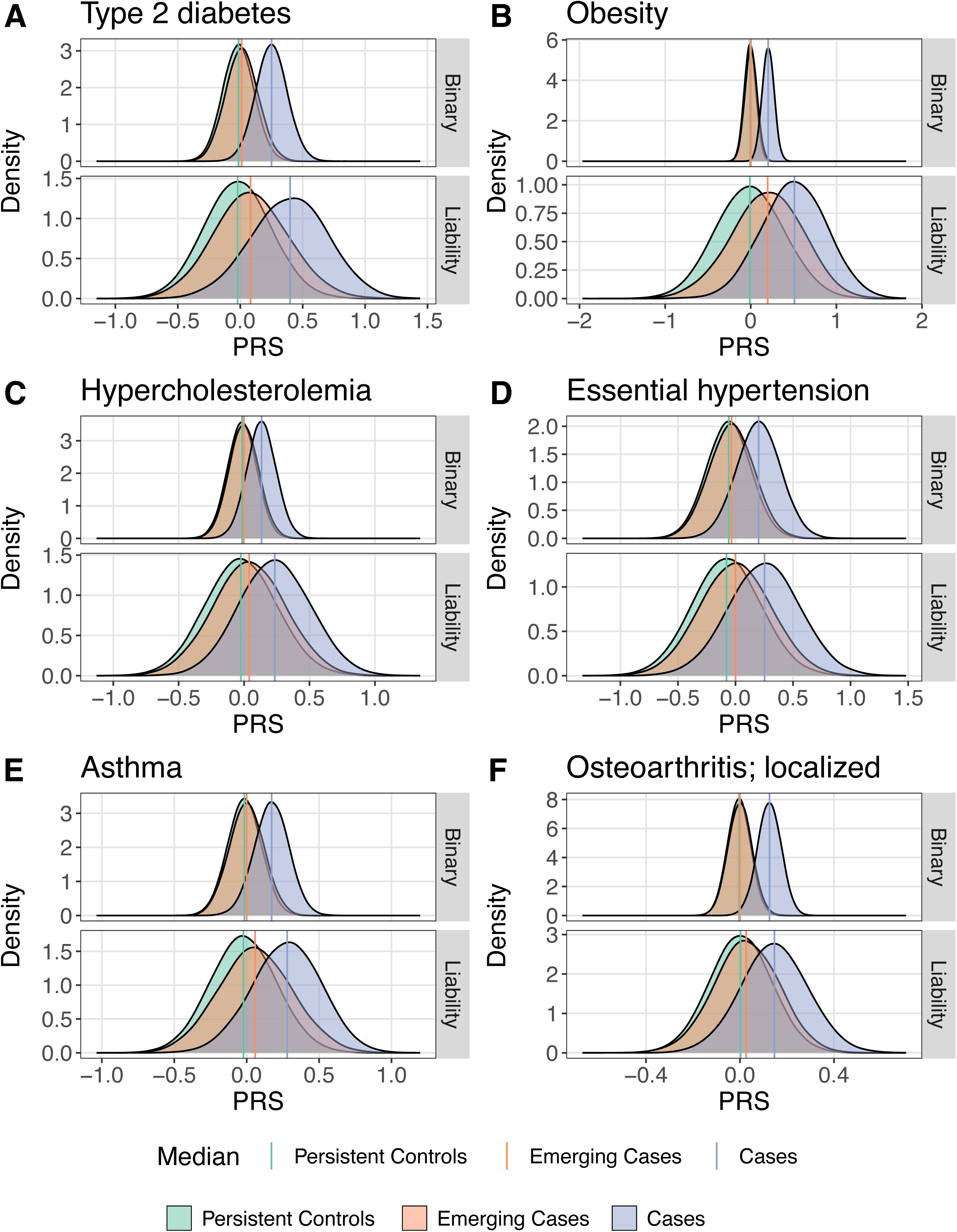
Comparison of PRS distributions between the binary and liability GWAS approaches. (A)-(F) PRS distributions for persistent controls (controls in the 2019 and 2022 release of the UKBB), emerging cases (controls in 2019 and cases in 2022) and cases (cases in 2019 and 2022) calculated through the binary and liability GWAS approaches across six diseases (type 2 diabetes, obesity, hypercholesterolemia, essential hypertension, asthma and localized osteoarthritis). Vertical lines denote the median value of the respective distribution.

**Figure 6.**
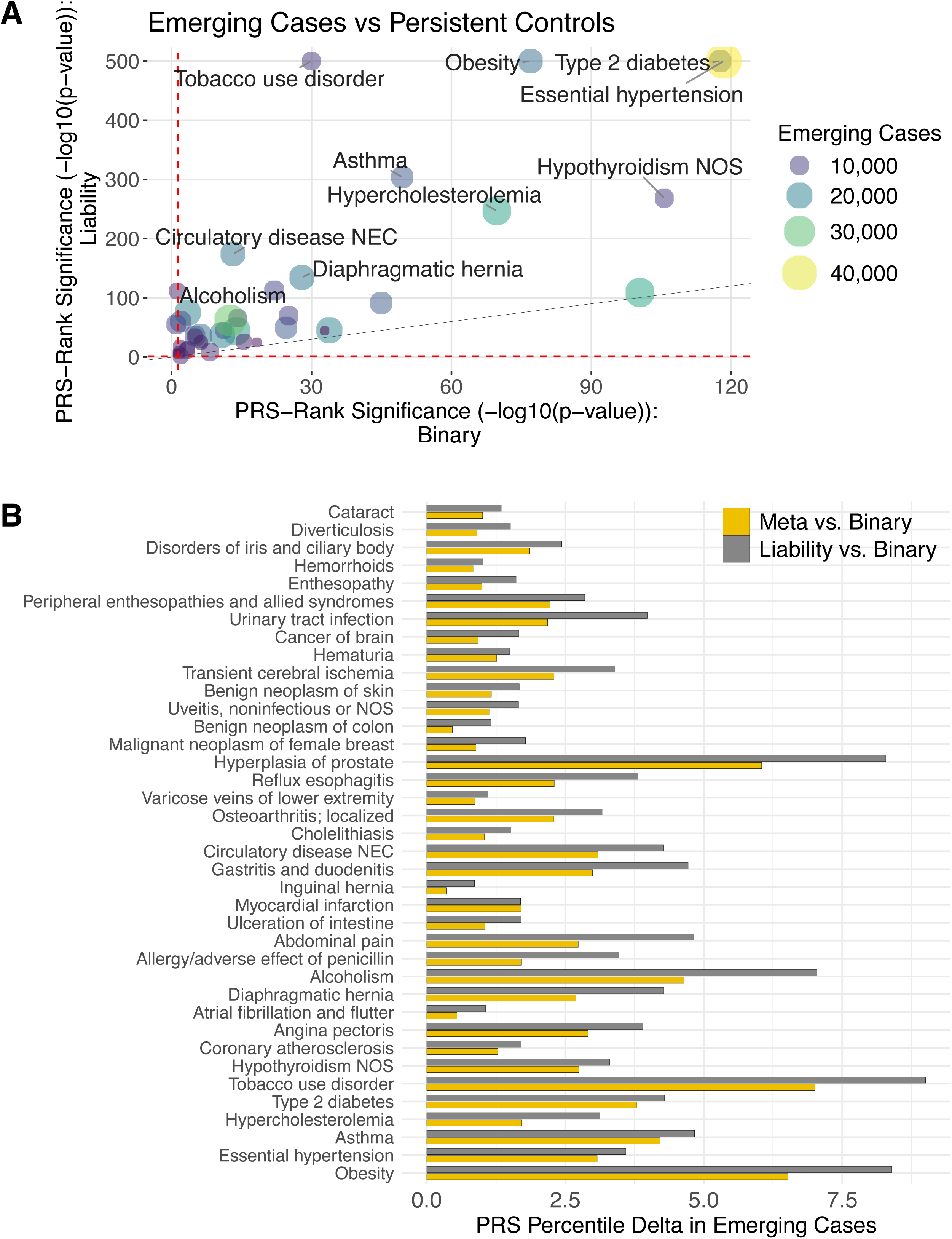
Difference in PRS between the binary, liability and meta GWAS approaches across traits. (A) Scatterplot comparing the difference in PRS between emerging cases and persistent controls for the binary (x-axis) and liability (y-axis) methods. The PRS difference results from two-sided Wilcoxon signed-rank tests and is plotted as a positive value if PRS allows to rank emerging cases higher than persistent controls. The size and color of the dots reflect the number of emerging cases (i.e., individuals who were controls in 2019 and cases in the 2022 UKBB release). (B) Difference in PRS percentile of emerging cases between liability and binary as well as between meta and binary methods. PRS percentiles were calculated relative to the entire dataset.

We additionally performed sensitivity analyses, estimating liability phenotypes and predictor effects in a distinct population (training set), thereby enabling independent testing in another population (test set). POPDx training was performed in 60% of the data selected at random, excluding the emerging cases, while GWAS was performed in the same 60% of the data, but including the emerging cases. In the test set (40% of the data), emerging cases still ranked significantly higher in liability-compared to binary-derived PRS (28/38 traits had a p-value *<* 0.05 in Wilcoxon rank tests, and 24/38 traits had a p-value *<* 0.05/38 in Wilcoxon rank tests) as well as in meta-compared to binary-derived PRS (24/38 traits had a p-value *<* 0.05/38 in Wilcoxon rank tests; Figures S6-8; Tables S17-19). Taken together, these analyses confirm that PRS derived by the liability and meta methods rank emerging cases higher than the binary method, thus holding a greater prediction value.

## Discussion

In this study, we presented and tested two new methods to conduct genetic associations which involve the modeling of disease liabilities from deep neural networks. We demonstrated how integrating a variety of patient data—including family history, socio-demographic characteristics, lifestyle surveys, physical measures, and laboratory examina-tions—can significantly contribute to boosting genetic association power and prediction performance. The deep neural networks were trained on the entire UKBB cohort to compute disease liability scores that align with the true diagnosis status and help impute missing diagnoses. The liability method resulted in higher counts of GWAS loci and improved PRS performance compared to using binary diagnosis values, while maintaining disease specificity. Furthermore, we presented the meta method which boosts performance by integrating liability phenotype scores of controls into the standard binary GWAS approach. Compared to the liability method, the genetic architecture of the meta method showed greater similarity to the binary approach, although it resulted in fewer novel GWAS counts and slightly lower performance in attributing high PRS to emerging cases. However, the meta method was found to be more robust when phenotype liability scores had low accuracies as demonstrated in the experiment with different feature sets. Thus, we recommend employing the liability method for diseases with high imputation accuracy and the meta method otherwise.

Computing GWAS on phenotype liability scores can potentially detect genetic associations that are not specific to the disease under investigation. When testing whether newly identified GWAS loci missed by the binary approach are significant in external larger GWAS, we found very high validation rates of 76% and 79% for the liability and meta method, respectively. The high validation rates provide reassurance that novel loci were indeed disease-specific. Additionally, in terms of absolute numbers, the liability method identified a greater number of loci with genome-wide significance in external GWAS compared to the binary method. Specifically, 365 loci significant in the liability method but missed by the binary method reached genome-wide significance, compared to 212 loci significant in the binary method but missed by the liability method. Likewise, genetic correlations with external GWAS were very similar between the binary and liability methods. Although genetic correlations were higher for the binary method, it should be noted that it has become common practice to include the UKBB in GWAS meta-analyses and thus the large majority of external GWAS included the UKBB and the standard CC classification. In the PRS analysis, we leveraged the fact that participant’s later disease diagnosis information was not available in the training data, making it an independent validation analysis for all three methods. The results indicated that PRS derived from the liability approaches outperformed the binary method in distinguishing emerging cases which held in sensitivity analyses where testing was conducted out-of-sample. Taken together, the findings suggest that incorporating liability phenotypes enhances genetic association studies while preserving a disease-relevant genetic architecture.

This study has several limitations. First, patient phenotyping is noisy, resulting in incorrect disease liabilities which in turn impairs the downstream genetic analysis. Second, our strategy is specific to the UKBB cohort; in addition to family history and medical data, our neural network model uses lifestyle, environmental exposures, and lab examinations to predict patient phenotypes. These data are not available in some observational studies. Third, for computational reasons, we limited the analysis to directly-genotyped SNPs which may limit the association strength in GWAS. We have not applied our approaches to other prospective cohort studies, however, the pipeline could be adapted to other biobanks. On a comparable scale, large population cohorts such as the “All of Us” program^41^, China Kadoorie Biobank^42^, and the US Million Veteran Program^43^ can be promising resources with more diverse populations to implement and replicate our genetic analyses. For all these cohorts, the assessment of disease liabilities may improve the power to detect genetic associations.

## Methods

### UK Biobank data

The UKBB is a cohort with rich phenotype data including the International Classification of Diseases, Tenth Revision (ICD-10) codes from which we could derive phecodes^4,44^. Phecodes are curated groups of ICD-10 codes for defining phenotypes^44,45^. We excluded individuals with no available ICD-10 codes in the 2019 data version and used the remaining white and unrelated individuals (N = 261,759) to infer phecodes and calculate disease liability scores (Section Modeling phenotype liabilities). Genetic data analyses were restricted to unrelated white British individuals that passed additional genetic quality control (QC) filters according to the UKBB (*used.in.pca.calculation* = 1 and *in.white.British.ancestry.subset* = 1 in Sample-QC v2 file)^4^. Furthermore, samples were excluded if the genetically inferred sex did not match the self-reported sex and if they had withdrawn their consent. Analyses were performed on autosomal genotype data provided by the UKBB. We filtered out SNPs with MAF below 1%, minor allele counts below 100, genotype missingess above 10% and Hardy-Weinberg equilibrium p-value exceeding 10e-15, resulting in a total of 589,086 SNP markers. All reported genetic analyses are for 261,759 unrelated white British individuals and sex-specific traits were analyzed on 142,817 females and 118,942 males, respectively.

### Phenotype data

ICD billing codes are commonly used to select individuals from large observational datasets such as the UKBB. Also, cases of multiple ICD codes are often put together as a group of patients to study a specific disease. Phecodes, combinations of relevant ICD codes used to describe phenotypes, have worked well in clinical research^45^. In this study, we retrieved 38 specific phecodes associated with more than 8,000 patients to represent 38 common diseases from the 2019 download of phenotype data. To obtain diagnostic labels for all the individuals in the UKBB, we mapped International Classification of Diseases Tenth Revision (ICD-10) codes to phecodes^44,46,47^. Furthermore, we retrieved an updated release of phenotype data (ICD-10 codes) from early 2022. From this data, we could distinguish between persistent controls (no disease diagnosis in 2019 and 2022) and emerging cases (disease diagnosis in 2022).

### Modelling phenotype liabilities

POPDx is an existing deep neural network model that outputs the phenotype liability scores for each UKBB participant^23^. We trained the POPDx framework on the phenotypic and health-related patient data. The framework processes the individuals’ data using a bilinear approach with two hidden layers of the POPDx architecture and a matrix transformation. The phecodes as gold standard labels are converted from ICD-10 diagnosis codes from UKBB. For the main results, we trained and tested POPDx on all individuals based on the 2019 version of phenotype data. We further conducted sensitivity analyses where the data was split into training (60%) and test sets (40%) for independent validation (see PRS sensitivity analysis with distinct training and test sets). To assess whether different feature sets might introduce non-disease-specific elements into the genetic architecture of diseases, we recomputed phenotype liability scores by 1) excluding features related to lifestyle, reducing the feature set from 39,469 to 26,085 features, and 2) computing phenotype liability scores solely on lifestyle features, which include 13,384 features. Lifestyle features include all those that fall into the categories of alcohol, alcohol use, cannabis use, diet, diet by 24-hour recall, education, electronic device use, employment, employment history, happiness and subjective well-being, household, residential air pollution, residential noise pollution, sleep, smoking, social support, sun exposure, traumatic events, unusual and psychotic experiences.

### GWAS calculation

We performed GWAS on all 38 complex traits following three different approaches: 1) logistic regression of case-control status (*binary*) using PLINK 2.0 with Firth fallback^48^, 2) quantitative GWAS on disease liability scores for all individuals (*liability*) using the LDAK software (v5.2, www.ldak.org^49^) and 3) meta-analyzing the binary GWAS (approach 1) with a quantitative GWAS on liability scores on controls only (*meta*) by applying inverse variance weighting^50^. As the covariance between liability scores on controls and case-control status of all individuals is zero, the two input GWAS of the meta-analysis are independent. Covariates used in all GWAS were sex, age, age^2^, and PCs 1–40. Sex was excluded from the covariates for traits specific to females and males. To reduce the computational burden, the analysis was performed on directly genotyped markers that passed quality control (see UK Biobank data).

### Defining GWAS loci

We performed clumping on SNPs passing the genome-wide significance threshold of 5e-8 to obtain independent SNP associations (*r*^2^ < 0.05 within a window of 1000kB). Clumping was performed using LDAK and as reference panel we used the genotypes of 20,000 randomly selected individuals among the included 261,807 samples. Next, we classified these independent signals into novel, common and missing signals in a pairwise analysis between the liability and binary method as well as between the meta and binary method. Novel signals are defined as signals found in the liability approaches, but missed in the binary method and missing signals are those present in the binary method, but missed in the liability approaches. To account for SNPs in linkage disequilibrium (LD) that have different significance levels across GWAS, we first extracted proxy SNPs of the clumped (tag) SNPs (*r*^2^ > 0.8, 1Mb window). A signal was classified as a novel if neither the tag SNP nor the proxy SNPs reached genome-wide significance in the binary GWAS. Likewise, a signal was classified as missing if neither the tag SNP of the binary GWAS nor the proxy SNPs reached genome-wide significance in the liability/meta GWAS. Tag/proxy SNPs significant in both GWAS were classified as common.

### Mapping SNPs to genes

We mapped the SNPs to genes using the reference annotation GENCODE GRCh37 release 42^51^. The GENCODE consortium provides public access to the most recent annotation of human reference genomes^51^. For each SNP, we searched through the genomic locations (genomic start and end positions) of all the protein-coding genes and non-coding RNA genes. SNPs needed to reside within the target genes or within a 100kb flanking region to be annotated.

### SNP-based heritability estimation

We calculated SNP heritabilities (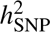) for each trait-method GWAS combination using the BLD-LDAK model^39^. BLD-LDAK estimates 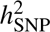 assuming that the expected 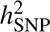 varies with LD and MAF while also taking into account 66 functional annotations. Estimates were calculated using SumHer (part of the LDAK software) and pre-computed tagging files designed for use with LDAK v5.2. Provided tagging files were computed on 2,000 white British individuals from the UKBB and we chose the set of non-ambiguous directly genotyped SNPs (MAF *>* 0.01, 577,525 markers of which 577,395 overlapped with our GWAS summmary statistics). For the binary method, heritabilities on the observed scale were transformed to the liability scale using the extension of the standard liability-scale model suitable for low-prevalence diseases where the population prevalence was set to the biobank sample prevalence^40^.

### Genetic correlations

We calculated genetic correlations (*r_g_*) using the LD score regression model implemented in LDSC v1.0.1 with pre-computed LD Scores from 1000 Genomes (European ancestry)^52,53^. First, we calculated pairwise genetic correlations between GWAS of binary and liability approaches (liability GWAS on controls, liability GWAS and meta GWAS). We repeated the analysis by computing phenotype liability scores with different feature sets (see Modelling phenotype liabilities) and re-calculated GWAS with these alternative scores. Second, we calculated pairwise genetic correlations with external GWAS.

### Polygenic risk score analysis

Per trait, we calculated PRS based on each of the three GWAS methods (binary, liability, meta) by using Quick PRS (LDAK v5.2) which relies on GWAS summary statistics^54^. Quick PRS requires a tagging file, heritability matrix and predictor-predictor correlations, of which pre-computed versions based on the UKBB data (computed using 2,000 white British individuals) are available on directly genotyped SNPs. Following the calculation of per-predictor heritabilities based on the GWAS summary statistics, tagging file and heritability matrix, per-predictor effect sizes (PRS effects) were estimated with the mega-prs command in a cross-validation (cv) fashion (recommended cv-proportion of 0.1) using the BayesR prediction model. From the per-predictor effect sizes, we estimated PRS for each individual by projecting their genetic data onto the estimated effect sizes.

Thus, each individual was assigned three scores, stemming from the binary, liability, and meta methods. To assess which method assigned higher PRS to disease cases we conducted two analyses, each involving emerging diseases (i.e., individuals that had a disease diagnosis in the 2022 UKBB release, but not in 2019). Information as to whether an individual turns into a case is not available to either method, and thus comparison at a later time point constitutes an independent validation analysis. In the first analysis, we compared which method better distinguishes between emerging cases and controls based on the PRS and conducted two-sided Wilcoxon signed-rank tests between these two groups within each method. In the second analysis, we focused only on emerging cases and tested which method assigned higher PRS in pairwise comparisons. To this end, we first ranked individuals relative to their PRS in the entire dataset. Each emerging case would thus have two ranks, one per method, and the difference in ranks was assessed in two-sided Wilcoxon signed-rank tests.

### PRS sensitivity analysis with distinct training and test sets

The POPDx model was trained exclusively on existing training data (60%) without emerging cases, while the GWAS included all the individuals in the training dataset. Based on these GWAS results, per-predictor effect sizes were estimated and PRS were computed on the remaining test data (40%).

## Data Availability

All data produced are available online at https://www.ukbiobank.ac.uk/enable-your-research/apply-for-access.

## Data and code availability

- This paper analyzes existing, publicly available data.
- All original code has been deposited at Github (https://github.com/luyang-ai4med/DL-GWAS).
- Any additional information about the analyses reported in this paper is available upon request.

## Acknowledgements

This work was supported by NIH GM102365, Agilent and the Chan-Zuckerberg Biohub. MCS was supported by the Swiss National Science Foundation (310030-189147) and would like to thank the Fulbright Program for funding her research stay at Stanford University.

The authors would like to thank Martin Jinye Zhang for the helpful discussion regarding heritability computations and gene annotations.

We would like to extend our gratitude to Doug Speed for his invaluable assistance in answering our questions about the LDAK software.

We would also like to thank Zoltán Kutalik for his contribution and helpful discussions regarding the definition of the meta approach.

## Author contributions

LY, MCS, and RBA conceived the original idea. LY and MCS carried out the experiments, and conducted the analyses. RBA supervised the project. All authors revised and approved the final manuscript.

## Supplemental Table legends

**Table S7** Genetic correlations with binary GWAS for different feature sets.

**Table S8** Genetic correlations with external GWAS.

**Table S20** Validation of novel loci identified by the liability method in external GWAS. Significance level of loci that reached genome-wide significance in the liability, but not binary GWAS in external GWAS of larger sample size.

**Table S21** Validation of novel loci identified by the meta method in external GWAS. Significance level of loci that reached genome-wide significance in the meta, but not binary GWAS in external GWAS of larger sample size.

