## Supplementary for "Genetic association studies using disease liabilities from deep neural networks"

### Contents

**1 Supplementary Note** **3**

**2 Supplementary Figures** **5**

**3 Supplementary Tables** **13**

S10 PRS of persistent controls, emerging cases, and cases calculated by the binary method. . 20
S11 PRS of persistent controls, emerging cases, and cases calculated by the liability method. 21
S12 PRS of persistent controls, emerging cases, and cases calculated by the meta method. . 22
S13 Difference in PRS rank between emerging cases and persistent controls for the binary, liability and meta methods . . . . . . 23
S14 PRS Percentile Delta in emerging Cases when comparing the binary, liability and meta methods. . . . . . 24

S15 PRS rank of emerging cases in binary, liability and meta methods. . . . . . . . . . . . . . . 25
S16 Difference in PRS rank of emerging cases between liability and binary methods as well as between meta and binary methods. . . . . . . . . . . . . . . . . . . . . . . . . . . . . . . . 26
S17 Difference in PRS rank between emerging cases and persistent controls for the binary, liability and meta methods calculated in an independent test set. . . . . . . . . . . . . . . . 27
S18 PRS rank of emerging cases in binary, liability and meta methods calculated in an independent test set. . . . . . . . . . . . . . . . . . . . . . . . . . . . . . . . . . . . . . . . 28
S19 Difference in PRS rank of emerging cases between liability and binary methods as well as between meta and binary methods calculated in an independent test set. . . . . . . . . . 29

### 1 Supplementary Note

The *meta* method consists of a meta-analysis between the binary GWAS and a quantitative GWAS on controls by using the disease liability scores. Thus, control individuals are used in both GWAS, however, the effective sample overlap is zero due to the null phenotypic correlation of these individuals in both analyses.

To show that the *meta* method does not suffer from genetic inflation, we conduct a simulation analysis. Briefly, we generate a disease with an underlying liability $Y$. This underlying liability is simulated to have a heritability $h^2$ caused by $m_c$ markers. Next, we classify the top 20% of the observed distribution $Y_{obs}$ as diseased individuals. On the full set of $m$ markers, a binary GWAS is then conducted on the defined cases and controls, a quantitative GWAS on $Y_{obs}$, but on controls only, and a meta-analysis between the two. Genomic inflation factors $\lambda_{GC}$ are calculated for all three GWAS.

**Simulation of genetic markers**

We simulated $m$ = 1,000 genetic markers for N = 50,000 individuals. Genotypes $G$ were generated from a binomial distribution with allele frequencies coming from a uniform distribution [0,1].

**Simulation of a disease liability**

Of the $m$ markers, $m_c$ = 100 causal markers were contributing to the heritability $h^2$ = 0.2. Genetic effect sizes $\beta_c$ for each causal marker $i$ were generated from a Gaussian under a model of negative selection where markers with smaller allele frequencies $q$ have larger effect sizes:

$$\beta_{c,i} \sim \mathcal{N}(0, \sigma^2[2q_i(1-q_i)]^{-0.25}) \tag{1}$$

From the generated heritability $H^2$ with $H^2 = \Sigma\beta_{c,i}^2 \cdot 2q_i(1-q_i)$, a scaling factor $f$ was derived ($f = \sqrt{h^2/H^2}$) to re-scale the effect sizes and to obtain the desired heritability $h^2$.

For each individual $j$, an observed disease liability $Y_{obs}$ was then simulated as follows:

$$Y_{obs,j} = \beta_{\mathbf{c}}{}^T \mathbf{G_{c,j}} + \epsilon_j \tag{2}$$

where $\epsilon_j$ comes from a normal distribution $\mathcal{N}(0,1)$.

**GWAS calculation**

1) A binary GWAS was conducted on cases and controls, where cases constituted the top 20% of the $Y_{obs}$ distribution.

2) A quantitative GWAS was conducted on the bottom 80% of the $Y_{obs}$ distribution, using $Y_{obs}$ as the outcome phenotype.

3) An inverse-weighted meta-analysis of these two GWAS was calculated using the effective sample size as weights.

##### Genomic inflation results

For each GWAS, we show the resulting QQ-plots and the genomic inflation factor $\lambda_{GC}$.

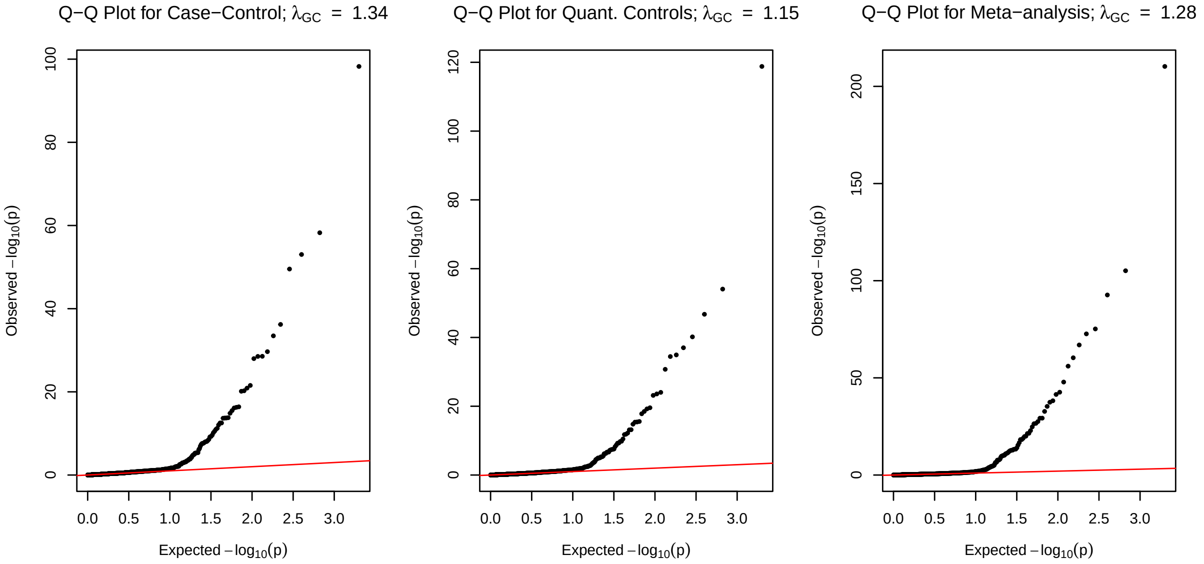

The genomic inflation factor for the binary GWAS, quantitative GWAS on controls, and meta-analysis was 1.34, 1.15 and 1.28, respectively, and no early deviation from the null distribution can be detected. If sample overlap were an issue for the *meta* approach, a strong deviation from the null distribution would be expected due to correlated errors. Together, these results demonstrate that the *meta* method does not suffer from genetic inflation, consistent with the zero phenotypic correlation among the overlapping control samples.

### 2 Supplementary Figures

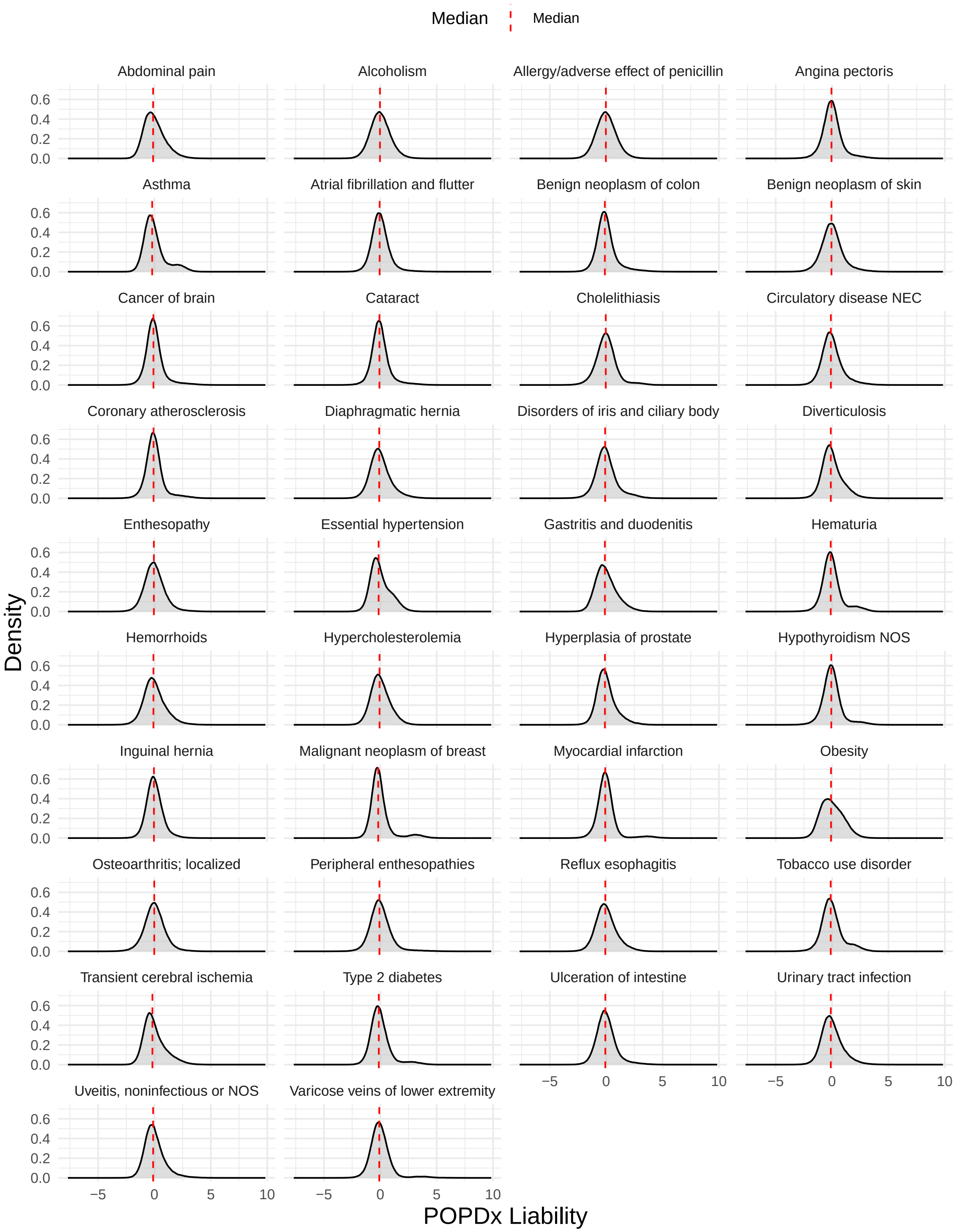

Figure S1: Phenotype liability scores computed by the POPDx deep neural network across the 38 diseases.

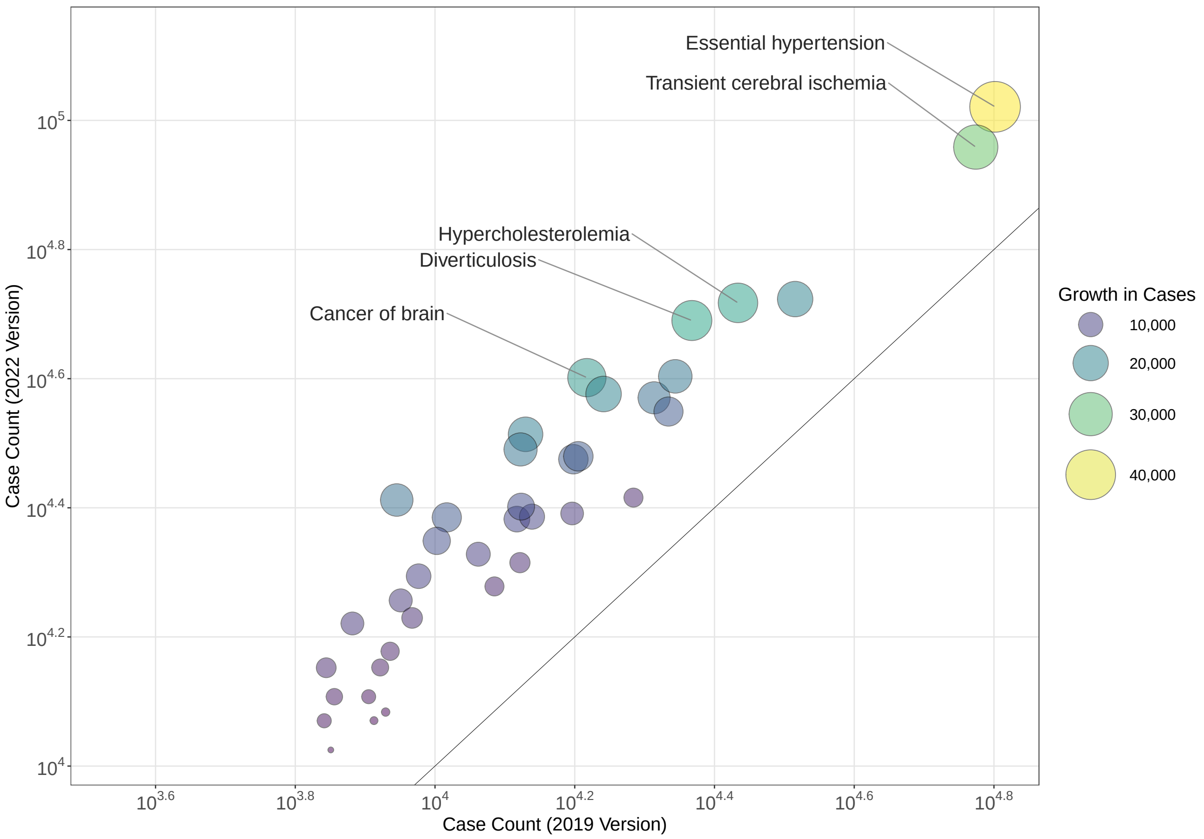

Figure S2: Number of cases per disease in the 2019 compared to the 2022 UK Biobank release.

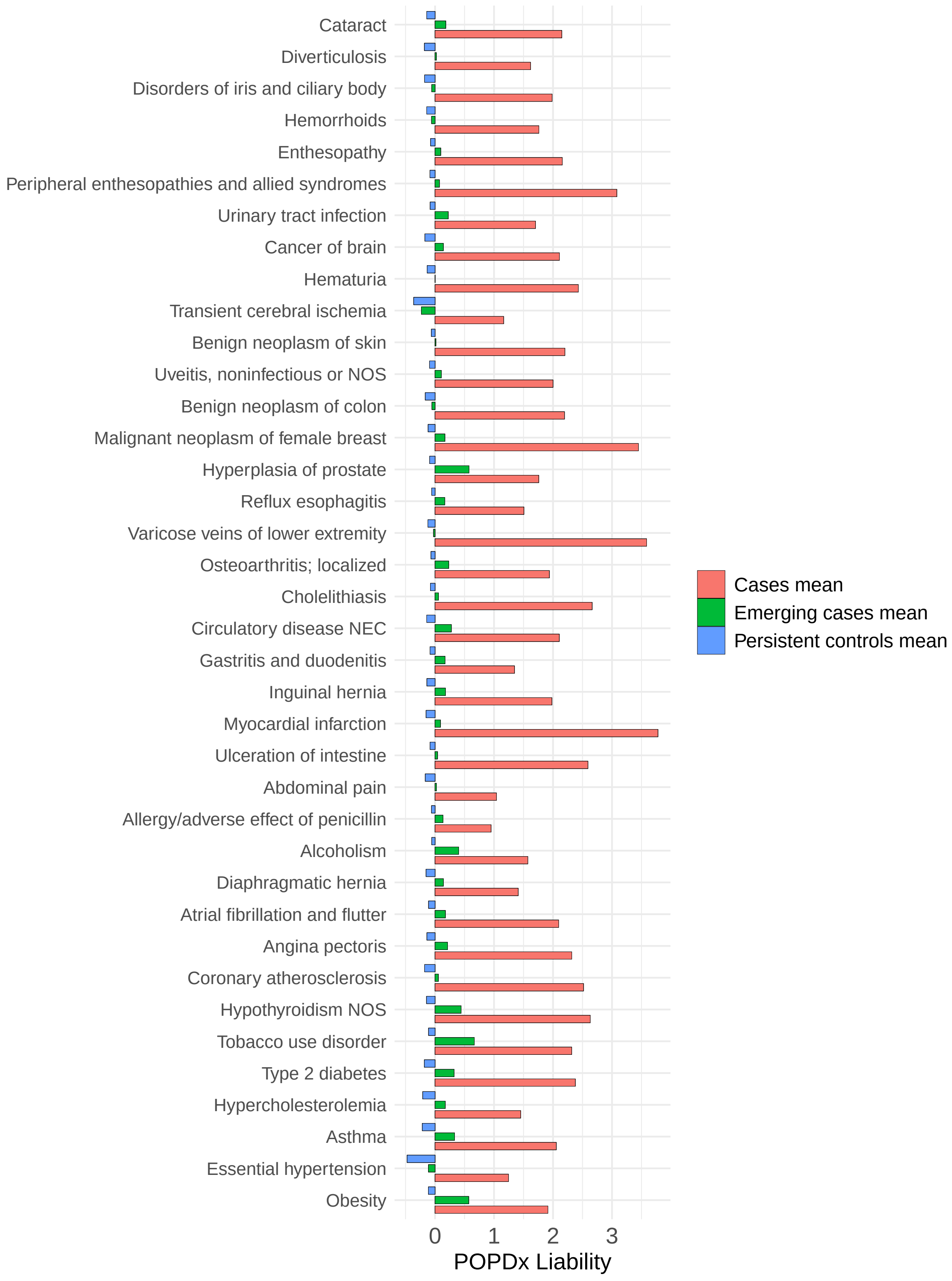

Figure S3: Disease liability scores of cases and emerging cases compared to persistent controls.

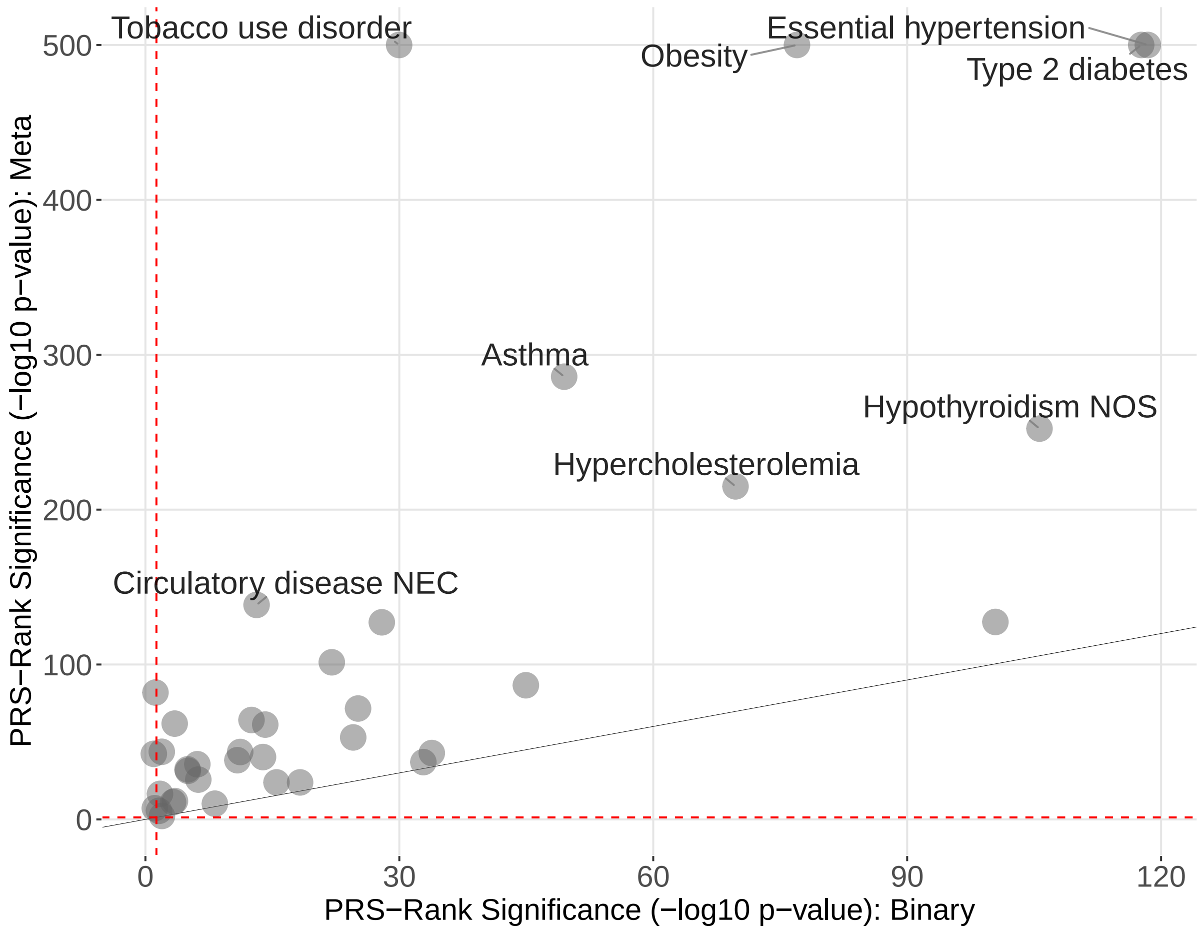

Figure S4: Scatterplot comparing the difference in PRS between emerging cases and persistent controls for the binary (x-axis) and meta (y-axis) methods. The PRS difference results from two-sided Wilcoxon signed-rank tests and is plotted as a positive value if PRS allow to rank emerging cases higher than persistent controls. P-values with numerical values below the computational limit were set to 10e-500.

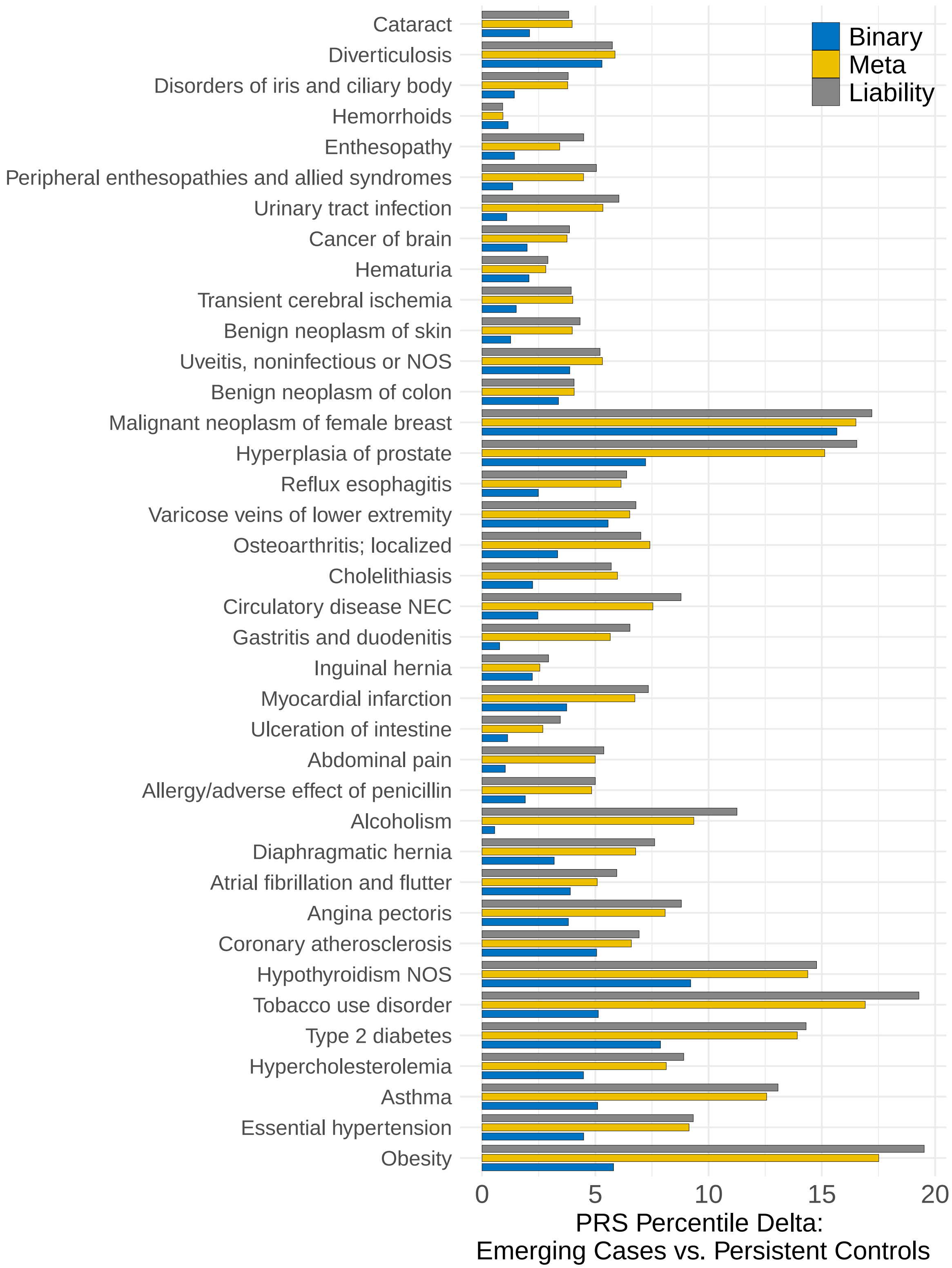

Figure S5: Difference in PRS percentiles between emerging cases and persistent controls calculated by the binary, liability and meta methods.

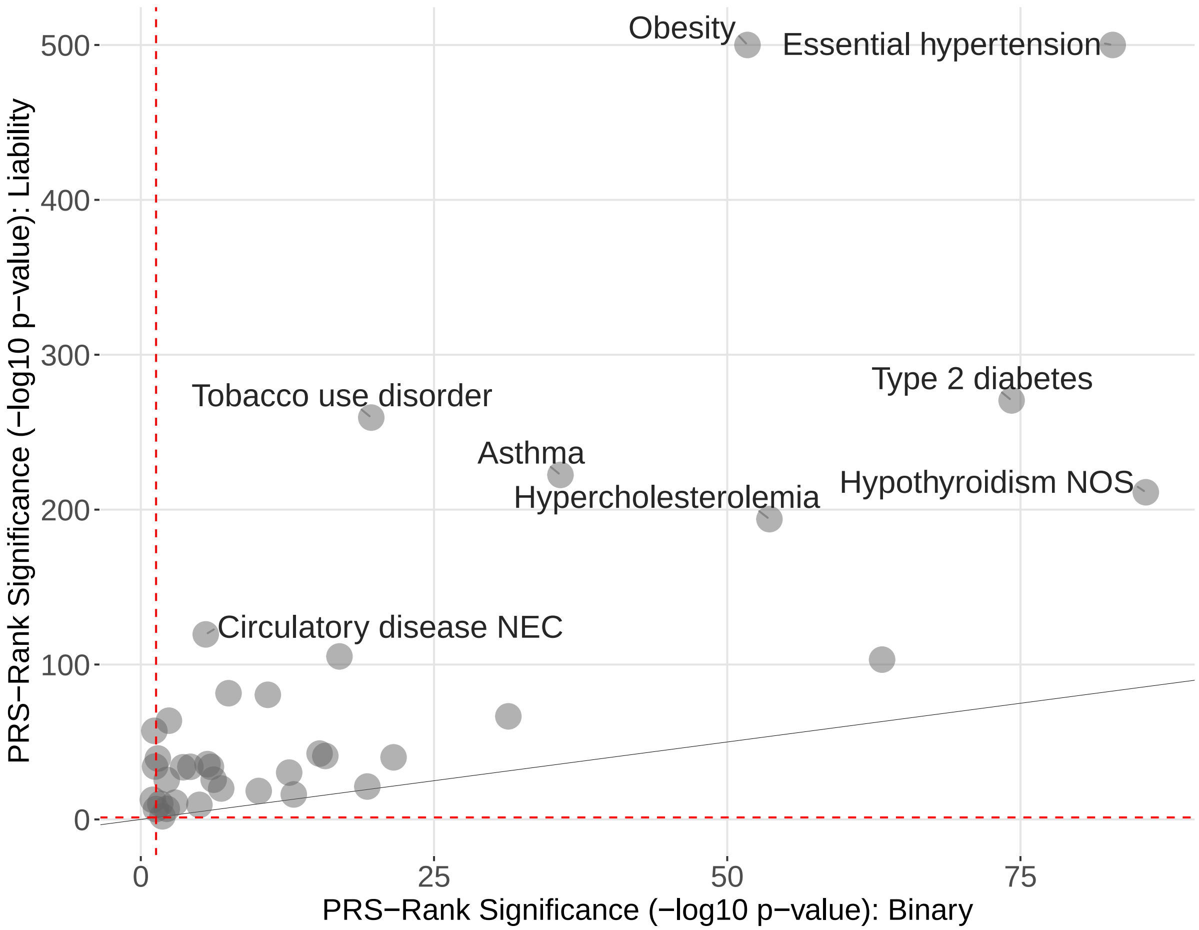

Figure S6: Scatterplot comparing the difference in PRS between emerging cases and persistent controls for the binary (x-axis) and liability (y-axis) methods in an independent test set selected at random. The PRS difference results from two-sided Wilcoxon signed-rank tests and is plotted as a positive value if PRS allow to rank emerging cases higher than persistent controls. P-values with numerical values below the computational limit were set to 10e-500.

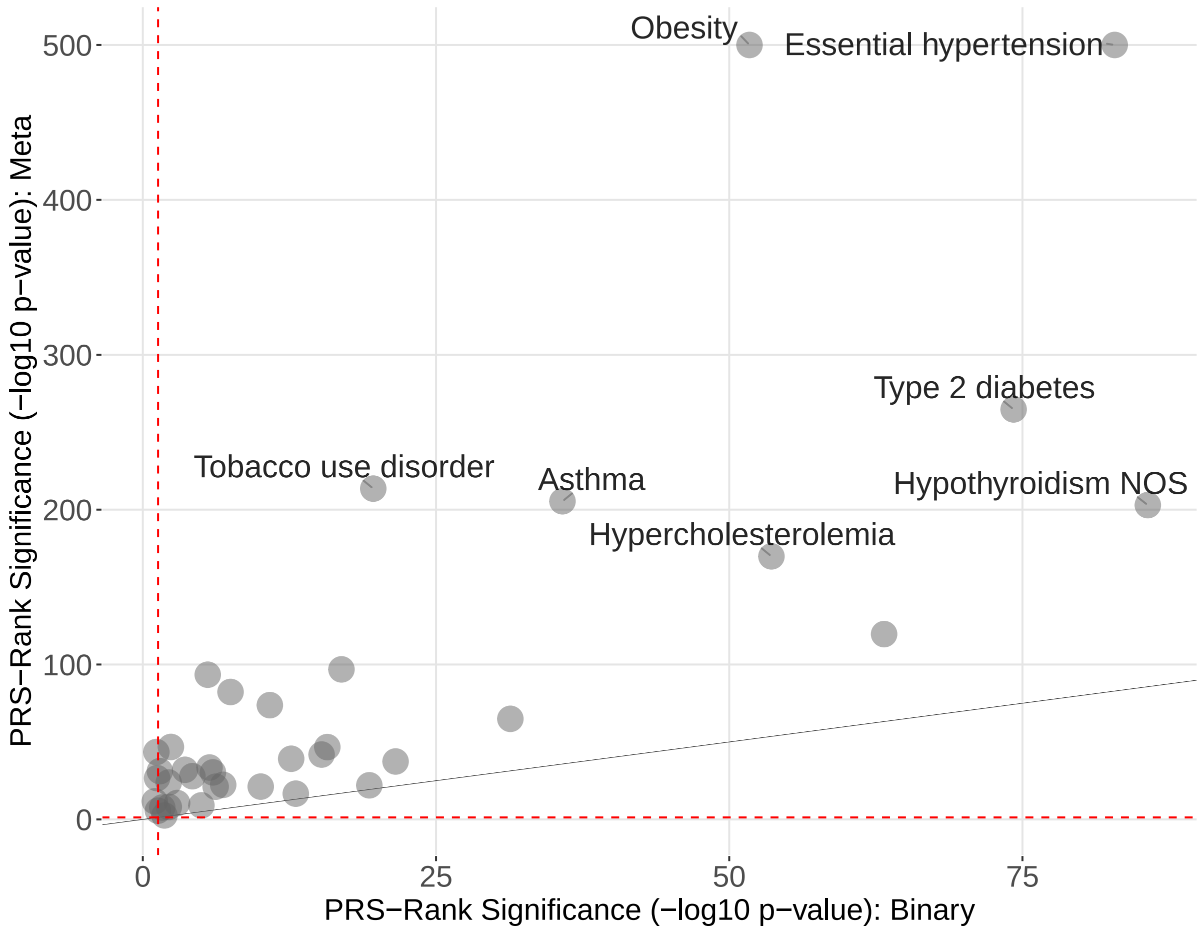

Figure S7: Scatterplot comparing the difference in PRS between emerging cases and persistent controls for the binary (x-axis) and meta (y-axis) methods in an independent test set selected at random. The PRS difference results from two-sided Wilcoxon signed-rank tests and is plotted as a positive value if PRS allow to rank emerging cases higher than persistent controls. P-values with numerical values below the computational limit were set to 10e-500.

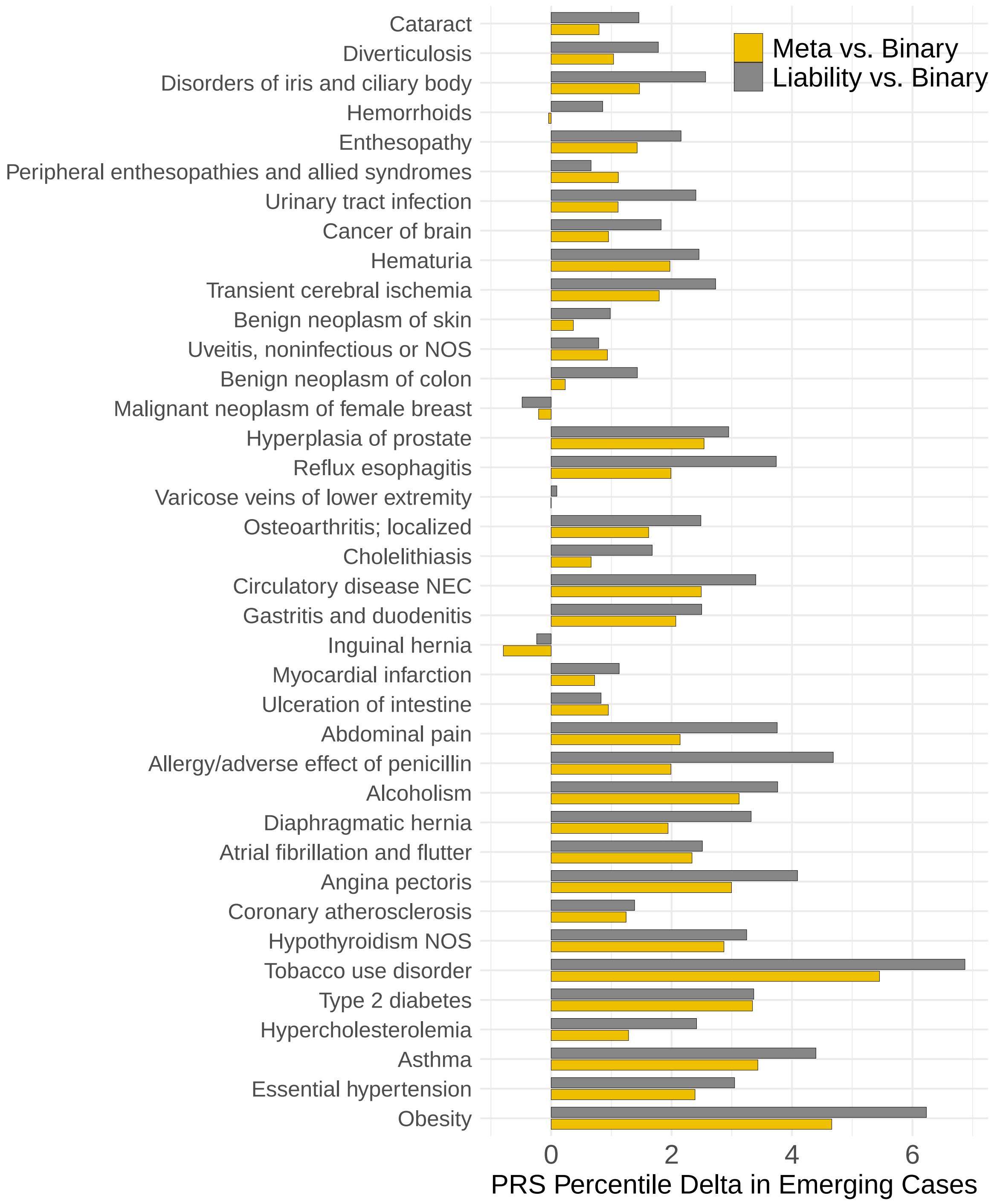

Figure S8: Difference in PRS percentile of emerging cases per trait between liability and binary as well as meta and binary methods in an independent test set selected at random. PRS percentiles were calculated relative to the entire dataset and the difference displayed here was calculated on newly diagnosed cases.

### 3 Supplementary Tables

| Disease/Trait | $h^2_{\text{SNP}}$(SE) (liability) | $h^2_{\text{SNP}}$(SE) (meta) | $h^2_{\text{SNP}}$(SE) (binary) | $h^2_{\text{SNP, liability}}$(SE) (binary) |
|---|---|---|---|---|
| Malignant neoplasm of female breast | 0.0512 (0.005) | 0.0262 (0.0026) | 0.0393 (0.0049) | 0.1595 (0.0259) |
| Cancer of brain | 0.0373 (0.0028) | 0.0213 (0.0014) | 0.0276 (0.0027) | 0.1062 (0.0132) |
| Benign neoplasm of colon | 0.0484 (0.0028) | 0.0264 (0.0016) | 0.0301 (0.0027) | 0.1116 (0.0129) |
| Benign neoplasm of skin | 0.0322 (0.0027) | 0.0131 (0.0014) | 0.0064 (0.0026) | 0.043 (0.0204) |
| Hypothyroidism NOS | 0.091 (0.003) | 0.0465 (0.0015) | 0.0557 (0.0028) | 0.2637 (0.0173) |
| Type 2 diabetes | 0.1899 (0.0034) | 0.0931 (0.0017) | 0.0694 (0.0029) | 0.2686 (0.0147) |
| Hypercholesterolemia | 0.1883 (0.0034) | 0.0897 (0.0018) | 0.0531 (0.0028) | 0.1511 (0.0108) |
| Obesity | 0.2848 (0.0038) | 0.1118 (0.0013) | 0.0372 (0.0028) | 0.2127 (0.0198) |
| Alcoholism | 0.2027 (0.0036) | 0.07 (0.0017) | 0.0105 (0.0026) | 0.0715 (0.021) |
| Tobacco use disorder | 0.1627 (0.0033) | 0.069 (0.0016) | 0.0266 (0.0027) | 0.1522 (0.0191) |
| Cataract | 0.0351 (0.0027) | 0.0187 (0.0014) | 0.022 (0.0027) | 0.0959 (0.0148) |
| Uveitis, noninfectious or NOS | 0.0358 (0.0027) | 0.019 (0.0014) | 0.0181 (0.0027) | 0.0935 (0.017) |
| Disorders of iris and ciliary body | 0.0375 (0.0028) | 0.0184 (0.0014) | 0.0127 (0.0026) | 0.0429 (0.0115) |
| Essential hypertension | 0.2204 (0.0036) | 0.1186 (0.002) | 0.1294 (0.0032) | 0.2414 (0.0068) |
| Myocardial infarction | 0.0582 (0.0028) | 0.0321 (0.0015) | 0.0327 (0.0027) | 0.1788 (0.0187) |
| Angina pectoris | 0.1269 (0.0032) | 0.0626 (0.0016) | 0.0426 (0.0028) | 0.1879 (0.0158) |
| Coronary atherosclerosis | 0.1001 (0.003) | 0.0551 (0.0016) | 0.0601 (0.0029) | 0.2317 (0.0144) |
| Atrial fibrillation and flutter | 0.0866 (0.003) | 0.0449 (0.0015) | 0.042 (0.0028) | 0.1942 (0.0164) |
| Transient cerebral ischemia | 0.0759 (0.0029) | 0.0433 (0.0017) | 0.0432 (0.0028) | 0.0834 (0.0068) |
| Varicose veins of lower extremity | 0.0564 (0.0028) | 0.0292 (0.0014) | 0.0481 (0.0028) | 0.2791 (0.0207) |
| Hemorrhoids | 0.0322 (0.0027) | 0.0184 (0.0014) | 0.0253 (0.0027) | 0.0886 (0.0122) |
| Circulatory disease NEC | 0.1029 (0.0031) | 0.0443 (0.0015) | 0.0166 (0.0027) | 0.0732 (0.0147) |
| Asthma | 0.1485 (0.0033) | 0.0737 (0.0017) | 0.0532 (0.0028) | 0.1724 (0.0122) |
| Reflux esophagitis | 0.1185 (0.0031) | 0.045 (0.0015) | 0.0091 (0.0026) | 0.0537 (0.0184) |
| Gastritis and duodenitis | 0.1183 (0.0031) | 0.0474 (0.0015) | 0.0128 (0.0026) | 0.0554 (0.0142) |
| Inguinal hernia | 0.0433 (0.0028) | 0.0238 (0.0014) | 0.0316 (0.0027) | 0.1249 (0.0138) |
| Diaphragmatic hernia | 0.1521 (0.0033) | 0.069 (0.0017) | 0.0413 (0.0028) | 0.1326 (0.0118) |
| Ulceration of intestine | 0.0862 (0.003) | 0.0394 (0.0015) | 0.0217 (0.0027) | 0.1334 (0.02) |
| Diverticulosis | 0.0498 (0.0028) | 0.0286 (0.0015) | 0.0495 (0.0028) | 0.1535 (0.0116) |
| Cholelithiasis | 0.0907 (0.003) | 0.038 (0.0015) | 0.0179 (0.0027) | 0.1192 (0.0212) |
| Urinary tract infection | 0.0689 (0.0029) | 0.0276 (0.0014) | 0.0045 (0.0026) | 0.0239 (0.0163) |
| Hematuria | 0.0295 (0.0027) | 0.0146 (0.0014) | 0.014 (0.0026) | 0.0619 (0.0146) |
| Hyperplasia of prostate | 0.0495 (0.0059) | 0.0292 (0.0031) | 0.0574 (0.0059) | 0.192 (0.0262) |
| Peripheral enthesopathies and allied syndromes | 0.0696 (0.0029) | 0.0314 (0.0014) | 0.0127 (0.0026) | 0.0865 (0.0212) |
| Enthesopathy | 0.0401 (0.0028) | 0.0178 (0.0014) | 0.0137 (0.0026) | 0.0823 (0.0189) |
| Osteoarthritis; localized | 0.0822 (0.003) | 0.0351 (0.0015) | 0.0217 (0.0027) | 0.1388 (0.0208) |
| Abdominal pain | 0.1251 (0.0032) | 0.0501 (0.0016) | 0.0219 (0.0027) | 0.0565 (0.0092) |
| Allergy/adverse effect of penicillin | 0.1727 (0.0034) | 0.0587 (0.0016) | 0.0142 (0.0026) | 0.0627 (0.0144) |

Table S1. SNP-based heritability ($h^2_{\text{SNP}}$) estimates calculated with the BLD-LDAK model. Heritability estimates for the binary method are reported both on the observed and transformed liability-scale.

| Disease/Trait | Study/Consortium | Reference | Sample Size N (Ncases/controls) | Meta-analysis with UK Biobank |
|---|---|---|---|---|
| Asthma | UK Biobank | Han et al., 2020 | 64,538 / 329,321 | Yes |
| Atrial fibrillation | GWAS meta-analysis | Nielsen et al., 2018 | 60,620 / 970,216 | Yes |
| BMI | GIANT & UK Biobank | Yengo et al., 2018 | 700,000 | Yes |
| Breast Cancer | GWAS meta-analysis | Michailidou et al., 2017 | 122,977 / 105,974 | No |
| Cataract | GWAS meta-analysis | Choquet et al., 2021 | 67,844 / 517,399 | Yes |
| Coronary Artery Disease | CARDIoGRAMplusC4D & UK Biobank | van der Harst et al., 2017 | 122,733 / 424,528 | Yes |
| Low-density lipoprotein cholesterol | GWAS meta-analysis | Graham et al., 2021 | 1,650,000 | Yes |
| Osteoarthritis | Genetics of Osteoarthritis consortium | Boer et al., 2021 | 177,517 / 649,173 | Yes |
| Stroke | MEGASTROKE Consortium | Malik et al., 2018 | 40,585 / 406,111 | No |
| Systolic blood pressure | UK Biobank & ICBP | Evangelou et al., 2018 | 757,601 | Yes |
| Type 2 diabetes | DIAGRAM Consortium & UK Biobank | Mahajan et al., 2018 | 74,124 / 824,006 | Yes |

Table S2. External GWAS used in the validation of novel SNPs and genetic correlations. These GWAS mostly stem from large meta-analyses, as well as from analyses conducted on quantitative phenotypes matching disease phenotypes. The last column indicates whether the UK Biobank was part of the GWAS meta-analysis.

| Disease/Trait | Threshold | | | | |
|---|---|---|---|---|---|
| | 0.5 | 0.6 | 0.7 | 0.8 | 0.9 |
| Angina pectoris | 0.9592 | 0.9473 | 0.9345 | 0.921 | 0.9067 |
| Cataract | 0.8955 | 0.8722 | 0.851 | 0.8273 | 0.8019 |
| Malignant neoplasm of female breast | 0.9979 | 0.9975 | 0.9966 | 0.9956 | 0.9936 |
| Benign neoplasm of skin | 0.9212 | 0.9047 | 0.8898 | 0.8737 | 0.8514 |
| Reflux esophagitis | 0.8067 | 0.7821 | 0.7546 | 0.7264 | 0.6955 |
| Hypothyroidism NOS | 0.9621 | 0.955 | 0.9469 | 0.9383 | 0.9293 |
| Essential hypertension | 0.8275 | 0.8022 | 0.7741 | 0.7423 | 0.7048 |
| Gastritis and duodenitis | 0.767 | 0.7406 | 0.7125 | 0.6809 | 0.6474 |
| Abdominal pain | 0.6714 | 0.6346 | 0.599 | 0.5625 | 0.5236 |
| Tobacco use disorder | 0.9569 | 0.9491 | 0.9399 | 0.9318 | 0.9201 |
| Myocardial infarction | 0.9991 | 0.9988 | 0.9988 | 0.9983 | 0.9983 |
| Osteoarthritis; localized | 0.8916 | 0.8746 | 0.8544 | 0.8289 | 0.8028 |
| Transient cerebral ischemia | 0.7365 | 0.703 | 0.6664 | 0.6285 | 0.5911 |
| Ulceration of intestine | 0.9513 | 0.9414 | 0.9327 | 0.9204 | 0.9082 |
| Diverticulosis | 0.8953 | 0.8729 | 0.8486 | 0.82 | 0.789 |
| Allergy/adverse effect of penicillin | 0.6632 | 0.623 | 0.5841 | 0.5454 | 0.5045 |
| Benign neoplasm of colon | 0.9186 | 0.9027 | 0.8858 | 0.8653 | 0.8454 |
| Alcoholism | 0.8437 | 0.8208 | 0.7936 | 0.7631 | 0.7289 |
| Hemorrhoids | 0.9082 | 0.8886 | 0.8669 | 0.8435 | 0.8166 |
| Obesity | 0.9604 | 0.951 | 0.939 | 0.9252 | 0.9062 |
| Hyperplasia of prostate | 0.9988 | 0.9981 | 0.9946 | 0.9888 | 0.9773 |
| Enthesopathy | 0.8995 | 0.8816 | 0.8627 | 0.8433 | 0.8233 |
| Asthma | 0.9147 | 0.9038 | 0.8924 | 0.8802 | 0.8688 |
| Coronary atherosclerosis | 0.9741 | 0.9677 | 0.9605 | 0.9505 | 0.9399 |
| Cancer of brain | 0.8817 | 0.8615 | 0.8367 | 0.811 | 0.7864 |
| Diaphragmatic hernia | 0.7643 | 0.7353 | 0.7058 | 0.6752 | 0.6443 |
| Varicose veins of lower extremity | 0.9866 | 0.9829 | 0.9808 | 0.9774 | 0.9755 |
| Circulatory disease NEC | 0.916 | 0.9001 | 0.8844 | 0.8652 | 0.8448 |
| Inguinal hernia | 0.9416 | 0.9275 | 0.9115 | 0.8919 | 0.872 |
| Hypercholesterolemia | 0.8813 | 0.8586 | 0.8325 | 0.8051 | 0.7739 |
| Type 2 diabetes | 0.9086 | 0.8899 | 0.8723 | 0.8526 | 0.8309 |
| Disorders of iris and ciliary body | 0.9333 | 0.9193 | 0.9038 | 0.8871 | 0.8683 |
| Hematuria | 0.9738 | 0.9683 | 0.9623 | 0.9564 | 0.9494 |
| Atrial fibrillation and flutter | 0.9134 | 0.8947 | 0.8709 | 0.8457 | 0.82 |
| Cholelithiasis | 0.9666 | 0.96 | 0.9539 | 0.9457 | 0.9366 |
| Peripheral enthesopathies and allied syndromes | 0.9631 | 0.9574 | 0.9509 | 0.9417 | 0.9332 |
| Urinary tract infection | 0.8421 | 0.8183 | 0.7891 | 0.7624 | 0.7348 |
| Uveitis, noninfectious or NOS | 0.8807 | 0.8611 | 0.8418 | 0.8215 | 0.799 |

Table S3. Sensitivity of POPDx for different thresholds.

| Disease/Trait | Threshold | | | | |
|---|---|---|---|---|---|
| | 0.5 | 0.6 | 0.7 | 0.8 | 0.9 |
| Angina pectoris | 0.7976 | 0.8344 | 0.8652 | 0.8914 | 0.9121 |
| Cataract | 0.7952 | 0.8316 | 0.8637 | 0.889 | 0.9101 |
| Malignant neoplasm of female breast | 0.7818 | 0.8239 | 0.86 | 0.8903 | 0.9151 |
| Benign neoplasm of skin | 0.7601 | 0.7937 | 0.8237 | 0.8501 | 0.8728 |
| Reflux esophagitis | 0.7467 | 0.7778 | 0.8059 | 0.8305 | 0.8526 |
| Hypothyroidism NOS | 0.8018 | 0.8387 | 0.87 | 0.8954 | 0.9164 |
| Essential hypertension | 0.8995 | 0.9178 | 0.9336 | 0.9475 | 0.9592 |
| Gastritis and duodenitis | 0.7549 | 0.7832 | 0.8087 | 0.8315 | 0.8524 |
| Abdominal pain | 0.7814 | 0.8094 | 0.8346 | 0.8573 | 0.8774 |
| Tobacco use disorder | 0.787 | 0.8161 | 0.8406 | 0.8611 | 0.8783 |
| Myocardial infarction | 0.8481 | 0.883 | 0.9119 | 0.9343 | 0.9514 |
| Osteoarthritis; localized | 0.7394 | 0.7754 | 0.8075 | 0.8363 | 0.8613 |
| Transient cerebral ischemia | 0.8839 | 0.9049 | 0.9234 | 0.9384 | 0.9511 |
| Ulceration of intestine | 0.7758 | 0.8099 | 0.8402 | 0.8647 | 0.8863 |
| Diverticulosis | 0.7993 | 0.8275 | 0.8521 | 0.8739 | 0.8931 |
| Allergy/adverse effect of penicillin | 0.7189 | 0.7523 | 0.7834 | 0.8121 | 0.8386 |
| Benign neoplasm of colon | 0.8282 | 0.8586 | 0.8846 | 0.9059 | 0.9235 |
| Alcoholism | 0.724 | 0.7569 | 0.7875 | 0.8152 | 0.8403 |
| Hemorrhoids | 0.7836 | 0.8132 | 0.8395 | 0.8635 | 0.8851 |
| Obesity | 0.7189 | 0.75 | 0.7788 | 0.8063 | 0.832 |
| Hyperplasia of prostate | 0.6131 | 0.6446 | 0.6855 | 0.7343 | 0.7859 |
| Enthesopathy | 0.7572 | 0.7903 | 0.8195 | 0.8458 | 0.8687 |
| Asthma | 0.8446 | 0.8676 | 0.8863 | 0.9017 | 0.9142 |
| Coronary atherosclerosis | 0.8379 | 0.8729 | 0.9015 | 0.9238 | 0.9416 |
| Cancer of brain | 0.8108 | 0.8457 | 0.8755 | 0.9 | 0.92 |
| Diaphragmatic hernia | 0.7861 | 0.8157 | 0.8418 | 0.8645 | 0.8843 |
| Varicose veins of lower extremity | 0.8109 | 0.8457 | 0.8761 | 0.9013 | 0.9226 |
| Circulatory disease NEC | 0.7905 | 0.8204 | 0.8464 | 0.8686 | 0.8884 |
| Inguinal hernia | 0.7613 | 0.797 | 0.8292 | 0.8577 | 0.8832 |
| Hypercholesterolemia | 0.7853 | 0.8141 | 0.8395 | 0.862 | 0.8825 |
| Type 2 diabetes | 0.8297 | 0.8602 | 0.8864 | 0.9082 | 0.9259 |
| Disorders of iris and ciliary body | 0.8164 | 0.8477 | 0.8747 | 0.8975 | 0.9166 |
| Hematuria | 0.8285 | 0.858 | 0.8815 | 0.9002 | 0.9155 |
| Atrial fibrillation and flutter | 0.7691 | 0.8044 | 0.8367 | 0.864 | 0.8876 |
| Cholelithiasis | 0.762 | 0.7993 | 0.8322 | 0.8612 | 0.8859 |
| Peripheral enthesopathies and allied syndromes | 0.7763 | 0.8101 | 0.8398 | 0.8659 | 0.8883 |
| Urinary tract infection | 0.7628 | 0.7913 | 0.8174 | 0.8409 | 0.8611 |
| Uveitis, noninfectious or NOS | 0.773 | 0.8033 | 0.83 | 0.8538 | 0.8747 |

Table S4. Specificity of POPDx for different thresholds.

| Disease/Trait | AUROC -All features | AUROC -All features except lifestyle | AUROC -Lifestyle |
|---|---|---|---|
| Myocardial infarction | 0.9995 | 0.9992 | 0.8165 |
| Malignant neoplasm of female breast | 0.9973 | 0.9969 | 0.8361 |
| Varicose veins of lower extremity | 0.9909 | 0.988 | 0.7228 |
| Coronary atherosclerosis | 0.9836 | 0.9751 | 0.8152 |
| Hematuria | 0.9761 | 0.9813 | 0.6443 |
| Hypothyroidism NOS | 0.9745 | 0.9752 | 0.7598 |
| Peripheral enthesopathies and allied syndromes | 0.9719 | 0.9634 | 0.6902 |
| Cholelithiasis | 0.9699 | 0.9569 | 0.7554 |
| Angina pectoris | 0.9682 | 0.9589 | 0.7989 |
| Hyperplasia of prostate | 0.9682 | 0.9578 | 0.8322 |
| Tobacco use disorder | 0.9608 | 0.9502 | 0.9504 |
| Ulceration of intestine | 0.9603 | 0.9621 | 0.8401 |
| Disorders of iris and ciliary body | 0.9563 | 0.9421 | 0.6717 |
| Inguinal hernia | 0.9519 | 0.9498 | 0.7835 |
| Benign neoplasm of colon | 0.9517 | 0.9587 | 0.7405 |
| Type 2 diabetes | 0.9511 | 0.9589 | 0.8425 |
| Asthma | 0.9511 | 0.9562 | 0.7275 |
| Obesity | 0.9439 | 0.9227 | 0.7834 |
| Essential hypertension | 0.9409 | 0.9393 | 0.7553 |
| Circulatory disease NEC | 0.9391 | 0.938 | 0.7343 |
| Cataract | 0.9351 | 0.9596 | 0.7501 |
| Atrial fibrillation and flutter | 0.9348 | 0.9301 | 0.7748 |
| Cancer of brain | 0.9332 | 0.9323 | 0.7152 |
| Benign neoplasm of skin | 0.933 | 0.948 | 0.7136 |
| Hemorrhoids | 0.9299 | 0.8831 | 0.6553 |
| Diverticulosis | 0.9248 | 0.9215 | 0.7354 |
| Enthesopathy | 0.9217 | 0.9434 | 0.7084 |
| Hypercholesterolemia | 0.9156 | 0.913 | 0.7594 |
| Osteoarthritis; localized | 0.9133 | 0.9089 | 0.7021 |
| Uveitis, noninfectious or NOS | 0.9132 | 0.9381 | 0.7108 |
| Transient cerebral ischemia | 0.8935 | 0.8605 | 0.6529 |
| Urinary tract infection | 0.8824 | 0.8986 | 0.6919 |
| Alcoholism | 0.8714 | 0.9319 | 0.8257 |
| Reflux esophagitis | 0.8591 | 0.8491 | 0.7181 |
| Diaphragmatic hernia | 0.8563 | 0.8767 | 0.74 |
| Gastritis and duodenitis | 0.8377 | 0.8606 | 0.7274 |
| Abdominal pain | 0.8042 | 0.8029 | 0.6422 |
| Allergy/adverse effect of penicillin | 0.7584 | 0.7904 | 0.7603 |

Table S5. AUROCs of POPDx with different feature sets.

| Disease/Trait | AUPRC -All features | AUPRC -All features except lifestyle | AUPRC -Lifestyle |
|---|---|---|---|
| Myocardial infarction | 0.996 | 0.991 | 0.1858 |
| Malignant neoplasm of female breast | 0.9601 | 0.9574 | 0.2143 |
| Varicose veins of lower extremity | 0.9463 | 0.9235 | 0.1059 |
| Coronary atherosclerosis | 0.8995 | 0.8586 | 0.2757 |
| Essential hypertension | 0.8659 | 0.8625 | 0.494 |
| Hypothyroidism NOS | 0.8332 | 0.8245 | 0.1399 |
| Peripheral enthesopathies and allied syndromes | 0.793 | 0.7394 | 0.0636 |
| Disorders of iris and ciliary body | 0.7866 | 0.7235 | 0.1603 |
| Angina pectoris | 0.7775 | 0.7385 | 0.2093 |
| Transient cerebral ischemia | 0.7717 | 0.7073 | 0.3546 |
| Hematuria | 0.7691 | 0.832 | 0.0866 |
| Asthma | 0.7584 | 0.784 | 0.2532 |
| Type 2 diabetes | 0.7527 | 0.7846 | 0.357 |
| Benign neoplasm of colon | 0.7487 | 0.7804 | 0.2024 |
| Inguinal hernia | 0.7196 | 0.7148 | 0.1946 |
| Cancer of brain | 0.7008 | 0.7005 | 0.1334 |
| Cholelithiasis | 0.6862 | 0.5803 | 0.1017 |
| Ulceration of intestine | 0.6757 | 0.6846 | 0.2348 |
| Cataract | 0.6709 | 0.7832 | 0.1519 |
| Hemorrhoids | 0.6525 | 0.4733 | 0.1414 |
| Diverticulosis | 0.6412 | 0.6288 | 0.2387 |
| Hyperplasia of prostate | 0.6373 | 0.5582 | 0.1697 |
| Atrial fibrillation and flutter | 0.6368 | 0.636 | 0.1602 |
| Hypercholesterolemia | 0.6309 | 0.6247 | 0.2759 |
| Circulatory disease NEC | 0.6251 | 0.6252 | 0.1436 |
| Tobacco use disorder | 0.6237 | 0.5841 | 0.5202 |
| Obesity | 0.53 | 0.4024 | 0.1404 |
| Enthesopathy | 0.5248 | 0.6327 | 0.0831 |
| Diaphragmatic hernia | 0.4905 | 0.5432 | 0.2343 |
| Osteoarthritis; localized | 0.4682 | 0.4501 | 0.0644 |
| Uveitis, noninfectious or NOS | 0.4644 | 0.555 | 0.0987 |
| Benign neoplasm of skin | 0.4624 | 0.5816 | 0.0941 |
| Abdominal pain | 0.4218 | 0.4214 | 0.2093 |
| Urinary tract infection | 0.3409 | 0.3968 | 0.0914 |
| Gastritis and duodenitis | 0.2928 | 0.3663 | 0.1497 |
| Alcoholism | 0.2678 | 0.5739 | 0.2056 |
| Reflux esophagitis | 0.2616 | 0.2423 | 0.104 |
| Allergy/adverse effect of penicillin | 0.1882 | 0.2664 | 0.2079 |

Table S6. AUPRCs of POPDx with different feature sets.

| Disease/Trait | Persistent controls mean (SD) | Emerging cases mean (SD) | Cases mean (SD) | T-test statistics | P-value |
|---|---|---|---|---|---|
| Angina pectoris | -0.138 (0.828) | 0.212 (0.967) | 2.318 (1.002) | 42.975 | 0.00E+00 |
| Cataract | -0.142 (0.835) | 0.182 (0.829) | 2.148 (1.258) | 51.515 | 0.00E+00 |
| Malignant neoplasm of female breast | -0.118 (0.782) | 0.167 (0.814) | 3.448 (0.764) | 25.178 | 1.03E-139 |
| Benign neoplasm of skin | -0.062 (0.92) | 0.014 (0.979) | 2.201 (1.219) | 4.868 | 1.13E-06 |
| Reflux esophagitis | -0.057 (0.954) | 0.165 (1.009) | 1.506 (1.093) | 18.422 | 9.76E-76 |
| Hypothyroidism NOS | -0.144 (0.795) | 0.442 (1.109) | 2.632 (0.972) | 70.096 | 0.00E+00 |
| Essential hypertension | -0.473 (0.663) | -0.112 (0.724) | 1.245 (0.765) | 96.933 | 0.00E+00 |
| Gastritis and duodenitis | -0.086 (0.935) | 0.17 (0.998) | 1.347 (1.099) | 27.442 | 1.53E-165 |
| Abdominal pain | -0.166 (0.885) | 0.025 (0.949) | 1.041 (1.087) | 29.024 | 7.11E-185 |
| Tobacco use disorder | -0.11 (0.877) | 0.666 (1.177) | 2.316 (0.874) | 81.803 | 0.00E+00 |
| Myocardial infarction | -0.152 (0.67) | 0.091 (0.752) | 3.778 (0.825) | 35.739 | 5.00E-279 |
| Osteoarthritis; localized | -0.069 (0.932) | 0.232 (0.976) | 1.938 (1.155) | 29.997 | 2.38E-197 |
| Transient cerebral ischemia | -0.362 (0.693) | -0.232 (0.715) | 1.163 (0.997) | 30.404 | 1.38E-202 |
| Ulceration of intestine | -0.084 (0.875) | 0.042 (0.915) | 2.591 (1.181) | 9.857 | 6.43E-23 |
| Diverticulosis | -0.18 (0.855) | 0.023 (0.863) | 1.619 (0.903) | 35.929 | 6.08E-282 |
| Allergy/adverse effect of penicillin | -0.06 (0.97) | 0.135 (0.974) | 0.95 (1.055) | 21.43 | 8.76E-102 |
| Benign neoplasm of colon | -0.166 (0.763) | -0.053 (0.795) | 2.194 (1.276) | 20.064 | 1.79E-89 |
| Alcoholism | -0.056 (0.956) | 0.4 (1.06) | 1.573 (1.07) | 39.825 | 0.00E+00 |
| Hemorrhoids | -0.142 (0.858) | -0.06 (0.885) | 1.761 (0.98) | 7.78 | 7.29E-15 |
| Obesity | -0.113 (0.919) | 0.571 (0.974) | 1.914 (0.777) | 93.244 | 0.00E+00 |
| Hyperplasia of prostate | -0.092 (0.947) | 0.576 (0.903) | 1.758 (0.444) | 70.304 | 0.00E+00 |
| Enthesopathy | -0.075 (0.904) | 0.097 (0.927) | 2.157 (1.305) | 14.377 | 7.50E-47 |
| Asthma | -0.216 (0.742) | 0.331 (1.041) | 2.055 (0.936) | 81.876 | 0.00E+00 |
| Coronary atherosclerosis | -0.178 (0.743) | 0.056 (0.781) | 2.517 (1.01) | 36.143 | 2.70E-285 |
| Cancer of brain | -0.172 (0.791) | 0.141 (0.785) | 2.109 (1.304) | 57.555 | 0.00E+00 |
| Diaphragmatic hernia | -0.152 (0.868) | 0.142 (0.939) | 1.409 (1.156) | 43.425 | 0.00E+00 |
| Varicose veins of lower extremity | -0.122 (0.731) | -0.026 (0.794) | 3.587 (1.242) | 7.858 | 3.93E-15 |
| Circulatory disease NEC | -0.142 (0.84) | 0.277 (0.976) | 2.106 (1.141) | 63.165 | 0.00E+00 |
| Inguinal hernia | -0.138 (0.86) | 0.177 (0.873) | 1.98 (0.923) | 33.901 | 2.47E-251 |
| Hypercholesterolemia | -0.209 (0.872) | 0.173 (0.878) | 1.452 (0.768) | 65.371 | 0.00E+00 |
| Type 2 diabetes | -0.181 (0.733) | 0.323 (1.011) | 2.378 (1.245) | 77.357 | 0.00E+00 |
| Disorders of iris and ciliary body | -0.178 (0.798) | -0.056 (0.824) | 1.985 (0.968) | 18.911 | 1.06E-79 |
| Hematuria | -0.133 (0.83) | 0.004 (0.885) | 2.429 (0.752) | 13.984 | 2.04E-44 |
| Atrial fibrillation and flutter | -0.11 (0.86) | 0.175 (0.919) | 2.096 (1.276) | 26.941 | 1.23E-159 |
| Cholelithiasis | -0.078 (0.891) | 0.058 (0.87) | 2.663 (1.023) | 11.338 | 8.69E-30 |
| Peripheral enthesopathies and allied syndromes | -0.087 (0.841) | 0.075 (0.892) | 3.081 (1.387) | 13.219 | 7.02E-40 |
| Urinary tract infection | -0.083 (0.919) | 0.223 (1.023) | 1.703 (1.226) | 35.743 | 4.36E-279 |
| Uveitis, noninfectious or NOS | -0.094 (0.889) | 0.107 (0.975) | 1.999 (1.287) | 25.763 | 3.62E-146 |

Table S9. Disease liability scores of persistent controls, emerging cases and cases per trait. P-values stem from a two-sided t-test comparing liability scores of persistent controls and emerging cases. Positive t-statistics indicate that emerging cases have higher PRS than persistent controls.

| Disease/Trait | Persistent controls PRS median (Binary) | Emerging cases PRS median (Binary) | Cases PRS median (Binary) | T-test statistics | P-value |
|---|---|---|---|---|---|
| Malignant neoplasm of female breast | -1.25E-02 | 7.34E-03 | 1.74E-01 | 12.3946 | 2.92E-35 |
| Cancer of brain | -7.96E-03 | -4.70E-03 | 1.08E-01 | 8.1163 | 4.83E-16 |
| Benign neoplasm of colon | -8.70E-03 | -2.48E-03 | 1.20E-01 | 12.4076 | 2.43E-35 |
| Benign neoplasm of skin | -8.44E-04 | 2.17E-04 | 7.65E-02 | 2.1023 | 3.55E-02 |
| Hypothyroidism NOS | -1.37E-02 | 1.89E-02 | 2.46E-01 | 22.751 | 1.83E-114 |
| Type 2 diabetes | -1.40E-02 | 1.18E-02 | 2.50E-01 | 23.9015 | 4.12E-126 |
| Hypercholesterolemia | -1.55E-02 | -2.12E-03 | 1.33E-01 | 18.4745 | 3.75E-76 |
| Obesity | -7.27E-03 | 3.03E-03 | 2.03E-01 | 19.2265 | 2.54E-82 |
| Alcoholism | -2.14E-03 | -1.71E-03 | 9.09E-02 | 2.0614 | 3.93E-02 |
| Tobacco use disorder | -4.62E-03 | 2.21E-03 | 1.43E-01 | 11.3673 | 6.19E-30 |
| Cataract | -5.72E-03 | -2.79E-03 | 9.60E-02 | 7.1922 | 6.39E-13 |
| Uveitis, noninfectious or NOS | -3.97E-03 | 1.24E-03 | 1.09E-01 | 10.2892 | 7.97E-25 |
| Disorders of iris and ciliary body | -4.83E-03 | -3.54E-03 | 5.81E-02 | 5.285 | 1.26E-07 |
| Essential hypertension | -5.85E-02 | -3.35E-02 | 2.02E-01 | 23.8414 | 1.87E-125 |
| Myocardial infarction | -5.63E-03 | 1.55E-03 | 1.66E-01 | 10.8761 | 1.52E-27 |
| Angina pectoris | -7.51E-03 | 3.76E-04 | 1.59E-01 | 9.9067 | 3.93E-23 |
| Coronary atherosclerosis | -1.35E-02 | 2.18E-03 | 2.10E-01 | 14.6511 | 1.39E-48 |
| Atrial fibrillation and flutter | -1.62E-02 | -4.60E-03 | 1.65E-01 | 8.0656 | 7.32E-16 |
| Transient cerebral ischemia | -1.92E-02 | -1.61E-02 | 7.94E-02 | 7.2596 | 3.90E-13 |
| Varicose veins of lower extremity | -8.86E-03 | 3.75E-03 | 2.45E-01 | 8.941 | 3.88E-19 |
| Hemorrhoids | -8.03E-03 | -6.46E-03 | 9.10E-02 | 2.258 | 2.39E-02 |
| Circulatory disease NEC | -5.62E-03 | -2.60E-03 | 7.71E-02 | 7.9516 | 1.85E-15 |
| Asthma | -1.56E-02 | -8.50E-05 | 1.72E-01 | 15.506 | 3.36E-54 |
| Reflux esophagitis | -1.78E-03 | 7.29E-04 | 9.34E-02 | 4.4717 | 7.76E-06 |
| Gastritis and duodenitis | -3.45E-03 | -2.76E-03 | 6.91E-02 | 1.5961 | 1.10E-01 |
| Inguinal hernia | -6.09E-03 | -1.93E-03 | 1.16E-01 | 5.8544 | 4.79E-09 |
| Diaphragmatic hernia | -7.16E-03 | -1.25E-03 | 1.31E-01 | 11.3361 | 8.84E-30 |
| Ulceration of intestine | -4.15E-03 | -2.66E-03 | 1.27E-01 | 2.3178 | 2.05E-02 |
| Diverticulosis | -1.15E-02 | 1.58E-03 | 1.51E-01 | 21.723 | 1.57E-104 |
| Cholelithiasis | -6.06E-03 | -2.63E-03 | 1.01E-01 | 6.0772 | 1.22E-09 |
| Urinary tract infection | -2.45E-03 | -1.82E-03 | 4.97E-02 | 2.8951 | 3.79E-03 |
| Hematuria | -3.17E-03 | -8.40E-04 | 6.58E-02 | 3.1982 | 1.38E-03 |
| Hyperplasia of prostate | -1.75E-02 | -1.05E-02 | 1.89E-01 | 6.8654 | 6.66E-12 |
| Peripheral enthesopathies and allied syndromes | -1.69E-03 | -5.14E-04 | 1.02E-01 | 2.6646 | 7.71E-03 |
| Enthesopathy | -9.76E-04 | 5.94E-04 | 8.33E-02 | 4.0243 | 5.72E-05 |
| Osteoarthritis; localized | -3.90E-03 | 3.53E-04 | 1.25E-01 | 8.119 | 4.72E-16 |
| Abdominal pain | -7.85E-03 | -6.53E-03 | 6.83E-02 | 3.5139 | 4.42E-04 |
| Allergy/adverse effect of penicillin | -5.38E-03 | -3.54E-03 | 7.66E-02 | 4.5642 | 5.02E-06 |

Table S10. PRS of persistent controls, emerging cases, and cases calculated by the binary method. P-values stem from a two-sided t-test comparing PRS of persistent controls and emerging cases. Positive t-statistics indicate that emerging cases have higher PRS than persistent controls.

| Disease/Trait | Persistent controls PRS median Liability | Emerging cases PRS median Liability | Cases PRS median Liability | T-test statistics | P-value |
|---|---|---|---|---|---|
| Malignant neoplasm of female breast | -9.40E-03 | 1.32E-02 | 1.64E-01 | 14.81 | 1.47E-49 |
| Cancer of brain | -5.20E-03 | 1.44E-03 | 7.55E-02 | 14.28 | 3.08E-46 |
| Benign neoplasm of colon | -6.78E-03 | 2.52E-03 | 1.05E-01 | 14.6 | 2.75E-48 |
| Benign neoplasm of skin | -9.34E-04 | 6.23E-03 | 7.07E-02 | 5.46 | 4.83E-08 |
| Hypothyroidism NOS | -1.19E-02 | 4.91E-02 | 2.40E-01 | 39.03 | 0.00E+00 |
| Type 2 diabetes | -2.07E-02 | 8.18E-02 | 3.99E-01 | 45.41 | 0.00E+00 |
| Hypercholesterolemia | -2.44E-02 | 3.88E-02 | 2.35E-01 | 34.33 | 1.43E-257 |
| Obesity | -1.11E-02 | 1.97E-01 | 5.10E-01 | 62.54 | 0.00E+00 |
| Alcoholism | -1.25E-02 | 6.81E-02 | 2.58E-01 | 23.71 | 4.06E-124 |
| Tobacco use disorder | -3.25E-03 | 1.25E-01 | 3.56E-01 | 46.77 | 0.00E+00 |
| Cataract | -4.86E-03 | 1.72E-03 | 7.84E-02 | 13.41 | 5.79E-41 |
| Uveitis, noninfectious or NOS | -5.45E-03 | 4.21E-03 | 7.08E-02 | 15.54 | 2.01E-54 |
| Disorders of iris and ciliary body | -6.20E-03 | 8.70E-04 | 7.10E-02 | 12.97 | 1.84E-38 |
| Essential hypertension | -7.74E-02 | 7.55E-04 | 2.52E-01 | 46.68 | 0.00E+00 |
| Myocardial infarction | -6.71E-03 | 1.38E-02 | 1.87E-01 | 18.54 | 1.16E-76 |
| Angina pectoris | -6.49E-03 | 3.74E-02 | 2.53E-01 | 24.04 | 1.67E-127 |
| Coronary atherosclerosis | -1.07E-02 | 1.88E-02 | 2.27E-01 | 21.02 | 5.48E-98 |
| Atrial fibrillation and flutter | -8.05E-03 | 1.43E-02 | 1.57E-01 | 10.95 | 6.40E-28 |
| Transient cerebral ischemia | -1.81E-02 | -5.51E-03 | 8.83E-02 | 17.02 | 6.28E-65 |
| Varicose veins of lower extremity | -6.21E-03 | 1.20E-02 | 1.92E-01 | 10.68 | 1.30E-26 |
| Hemorrhoids | -1.48E-03 | 6.00E-06 | 6.44E-02 | 2.7 | 6.86E-03 |
| Circulatory disease NEC | -1.66E-03 | 3.51E-02 | 1.88E-01 | 29.64 | 1.14E-192 |
| Asthma | -2.22E-02 | 5.83E-02 | 2.80E-01 | 40.43 | 0.00E+00 |
| Reflux esophagitis | 1.55E-03 | 3.23E-02 | 1.65E-01 | 13.17 | 1.27E-39 |
| Gastritis and duodenitis | -4.94E-04 | 2.95E-02 | 1.50E-01 | 16.05 | 6.41E-58 |
| Inguinal hernia | -4.69E-04 | 5.45E-03 | 7.95E-02 | 6.3 | 3.03E-10 |
| Diaphragmatic hernia | -2.32E-03 | 4.26E-02 | 2.03E-01 | 25.96 | 2.25E-148 |
| Ulceration of intestine | -4.13E-03 | 7.62E-03 | 1.94E-01 | 5.68 | 1.34E-08 |
| Diverticulosis | -6.02E-03 | 7.83E-03 | 8.36E-02 | 22.96 | 1.49E-116 |
| Cholelithiasis | 3.80E-03 | 2.50E-02 | 2.24E-01 | 10.42 | 2.02E-25 |
| Urinary tract infection | 1.24E-03 | 2.06E-02 | 1.14E-01 | 17.17 | 4.97E-66 |
| Hematuria | -3.15E-03 | 1.34E-03 | 6.39E-02 | 7.18 | 7.01E-13 |
| Hyperplasia of prostate | -9.54E-03 | 7.13E-03 | 9.96E-02 | 14.66 | 1.28E-48 |
| Peripheral enthesopathies and allied syndromes | 2.03E-03 | 1.76E-02 | 2.03E-01 | 9.26 | 2.03E-20 |
| Enthesopathy | 8.11E-04 | 8.67E-03 | 7.78E-02 | 7.84 | 4.53E-15 |
| Osteoarthritis; localized | 2.45E-03 | 2.63E-02 | 1.47E-01 | 17.41 | 7.61E-68 |
| Abdominal pain | -5.97E-03 | 2.07E-02 | 1.30E-01 | 19.04 | 9.32E-81 |
| Allergy/adverse effect of penicillin | 2.12E-03 | 3.40E-02 | 1.62E-01 | 12.61 | 2.03E-36 |

Table S11. PRS of persistent controls, emerging cases, and cases calculated by the liability method. P-values stem from a two-sided t-test comparing PRS of persistent controls and emerging cases. Positive t-statistics indicate that emerging cases have higher PRS than persistent controls.

| Disease/Trait | Persistent controls PRS median Meta | Emerging cases PRS median Meta | Cases PRS median Meta | T-test statistics | P-value |
|---|---|---|---|---|---|
| Malignant neoplasm of female breast | -6.38E-03 | 8.11E-03 | 1.11E-01 | 13.7 | 1.08E-42 |
| Cancer of brain | -5.37E-03 | -2.30E-04 | 7.72E-02 | 13.85 | 1.34E-43 |
| Benign neoplasm of colon | -6.01E-03 | 9.46E-04 | 9.50E-02 | 13.94 | 3.71E-44 |
| Benign neoplasm of skin | -1.67E-03 | 2.50E-03 | 7.98E-02 | 5.47 | 4.63E-08 |
| Hypothyroidism NOS | -1.03E-02 | 3.37E-02 | 2.02E-01 | 37.37 | 8.03E-305 |
| Type 2 diabetes | -1.61E-02 | 5.33E-02 | 3.10E-01 | 45.1 | 0.00E+00 |
| Hypercholesterolemia | -2.11E-02 | 1.85E-02 | 2.15E-01 | 32.11 | 1.10E-225 |
| Obesity | -1.28E-02 | 9.63E-02 | 4.79E-01 | 56.88 | 0.00E+00 |
| Alcoholism | -1.03E-02 | 2.21E-02 | 3.25E-01 | 20.06 | 1.90E-89 |
| Tobacco use disorder | -5.22E-03 | 6.13E-02 | 3.04E-01 | 41.45 | 0.00E+00 |
| Cataract | -4.77E-03 | 3.70E-04 | 7.87E-02 | 13.58 | 5.09E-42 |
| Uveitis, noninfectious or NOS | -5.21E-03 | 2.16E-03 | 9.14E-02 | 15.97 | 2.07E-57 |
| Disorders of iris and ciliary body | -4.83E-03 | 1.55E-04 | 6.01E-02 | 12.7 | 6.30E-37 |
| Essential hypertension | -5.93E-02 | -2.78E-03 | 1.94E-01 | 46.7 | 0.00E+00 |
| Myocardial infarction | -4.22E-03 | 9.72E-03 | 1.45E-01 | 18.74 | 2.57E-78 |
| Angina pectoris | -7.52E-03 | 2.03E-02 | 2.21E-01 | 22.55 | 1.90E-112 |
| Coronary atherosclerosis | -9.09E-03 | 1.18E-02 | 1.87E-01 | 20.45 | 7.50E-93 |
| Atrial fibrillation and flutter | -9.18E-03 | 5.01E-03 | 1.72E-01 | 10.52 | 7.40E-26 |
| Transient cerebral ischemia | -1.76E-02 | -7.76E-03 | 7.60E-02 | 17.4 | 8.48E-68 |
| Varicose veins of lower extremity | -4.54E-03 | 8.10E-03 | 1.48E-01 | 10.46 | 1.38E-25 |
| Hemorrhoids | -2.88E-03 | -1.71E-03 | 6.87E-02 | 2.99 | 2.84E-03 |
| Circulatory disease NEC | -4.38E-03 | 1.50E-02 | 1.59E-01 | 26.41 | 1.53E-153 |
| Asthma | -1.73E-02 | 3.69E-02 | 2.13E-01 | 39.07 | 0.00E+00 |
| Reflux esophagitis | -1.41E-03 | 1.47E-02 | 2.04E-01 | 12.47 | 1.14E-35 |
| Gastritis and duodenitis | -4.97E-03 | 1.02E-02 | 1.70E-01 | 13.68 | 1.34E-42 |
| Inguinal hernia | -2.00E-03 | 2.10E-03 | 8.82E-02 | 6.58 | 4.57E-11 |
| Diaphragmatic hernia | -7.51E-03 | 1.82E-02 | 1.97E-01 | 25.19 | 7.41E-140 |
| Ulceration of intestine | -4.59E-03 | 1.47E-03 | 1.82E-01 | 4.84 | 1.28E-06 |
| Diverticulosis | -6.92E-03 | 3.86E-03 | 9.03E-02 | 25.03 | 4.49E-138 |
| Cholelithiasis | -7.60E-04 | 1.30E-02 | 1.95E-01 | 10.66 | 1.55E-26 |
| Urinary tract infection | -3.47E-03 | 6.19E-03 | 1.24E-01 | 14.49 | 1.38E-47 |
| Hematuria | -2.65E-03 | 3.81E-04 | 5.45E-02 | 7.04 | 1.93E-12 |
| Hyperplasia of prostate | -9.92E-03 | 2.33E-03 | 1.11E-01 | 14.47 | 2.04E-47 |
| Peripheral enthesopathies and allied syndromes | -2.53E-04 | 8.49E-03 | 1.75E-01 | 8.84 | 9.62E-19 |
| Enthesopathy | 2.70E-05 | 4.01E-03 | 8.50E-02 | 7.26 | 3.85E-13 |
| Osteoarthritis; localized | -2.58E-03 | 1.27E-02 | 1.72E-01 | 16.7 | 1.48E-62 |
| Abdominal pain | -1.01E-02 | 4.28E-03 | 1.20E-01 | 16.83 | 1.59E-63 |
| Allergy/adverse effect of penicillin | -8.64E-03 | 6.68E-03 | 2.09E-01 | 11.82 | 3.07E-32 |

Table S12. PRS of persistent controls, emerging cases, and cases calculated by the meta method. P-values stem from a two-sided t-test comparing PRS of persistent controls and emerging cases. Positive t-statistics indicate that emerging cases have higher PRS than persistent controls.

| Disease/Trait | Wilcoxon statistic (Binary) | P-value (Binary) | Wilcoxon statistic (Liability) | P-value (Liability) | Wilcoxon statistic (Meta) | P-value (Meta) |
|---|---|---|---|---|---|---|
| Abdominal pain | 3.8 | 3.48E-04 | 20.07 | 1.33E-89 | 17.68 | 5.92E-70 |
| Alcoholism | 1.85 | 6.48E-02 | 21.85 | 7.18E-106 | 18.68 | 6.97E-78 |
| Allergy/adverse effect of penicillin | 4.43 | 9.64E-06 | 16.05 | 5.45E-58 | 14.46 | 2.25E-47 |
| Angina pectoris | 9.82 | 9.56E-23 | 24.21 | 1.62E-129 | 23.14 | 1.76E-118 |
| Asthma | 14.90 | 3.35E-50 | 38.44 | 0.00E+00 | 36.18 | 1.35E-286 |
| Atrial fibrillation and flutter | 8.16 | 3.24E-16 | 9.21 | 3.27E-20 | 9.75 | 1.84E-22 |
| Benign neoplasm of colon | 12.26 | 1.46E-34 | 15.15 | 8.00E-52 | 15.20 | 3.60E-52 |
| Benign neoplasm of skin | 1.76 | 7.79E-02 | 4.86 | 1.17E-06 | 5.55 | 2.81E-08 |
| Cancer of brain | 7.71 | 1.28E-14 | 14.38 | 7.33E-47 | 14.02 | 1.10E-44 |
| Cataract | 6.76 | 1.36E-11 | 15.46 | 6.19E-54 | 14.81 | 1.36E-49 |
| Cholelithiasis | 5.00 | 5.63E-07 | 10.86 | 1.75E-27 | 10.84 | 2.23E-27 |
| Circulatory disease NEC | 7.48 | 7.35E-14 | 30.42 | 2.90E-203 | 27.16 | 2.18E-162 |
| Coronary atherosclerosis | 14.19 | 1.12E-45 | 21.45 | 5.07E-102 | 20.83 | 2.44E-96 |
| Diaphragmatic hernia | 11.11 | 1.18E-28 | 26.68 | 7.06E-157 | 25.76 | 2.77E-146 |
| Disorders of iris and ciliary body | 4.95 | 7.31E-07 | 16.38 | 2.68E-60 | 15.77 | 4.67E-56 |
| Diverticulosis | 21.35 | 3.76E-101 | 25.96 | 1.25E-148 | 27.91 | 2.03E-171 |
| Enthesopathy | 3.60 | 3.23E-04 | 7.35 | 2.03E-13 | 6.97 | 3.24E-12 |
| Essential hypertension | 23.21 | 3.68E-119 | 51.90 | 0.00E+00 | 51.19 | 0.00E+00 |
| Gastritis and duodenitis | 1.64 | 1.02E-01 | 16.08 | 3.76E-58 | 13.86 | 1.06E-43 |
| Hematuria | 3.46 | 5.41E-04 | 6.91 | 5.01E-12 | 6.97 | 3.27E-12 |
| Hemorrhoids | 2.54 | 1.12E-02 | 3.48 | 5.07E-04 | 3.82 | 1.32E-04 |
| Hypercholesterolemia | 17.74 | 1.91E-70 | 34.86 | 3.44E-266 | 32.79 | 9.47E-236 |
| Hyperplasia of prostate | 6.87 | 6.33E-12 | 15.79 | 3.59E-56 | 15.13 | 1.05E-51 |
| Hypothyroidism NOS | 21.91 | 2.31E-106 | 39.3 | 0.00E+00 | 37.23 | 1.95E-303 |
| Inguinal hernia | 5.81 | 6.25E-09 | 7.83 | 4.94E-15 | 7.60 | 3.04E-14 |
| Malignant neoplasm of female breast | 12.07 | 1.45E-33 | 12.84 | 1.01E-37 | 12.78 | 2.00E-37 |
| Myocardial infarction | 10.51 | 7.50E-26 | 17.06 | 2.96E-65 | 17.92 | 7.89E-72 |
| Obesity | 18.66 | 1.04E-77 | 61.72 | 0.00E+00 | 56.30 | 0.00E+00 |
| Osteoarthritis; localized | 7.79 | 6.94E-15 | 17.59 | 2.81E-69 | 1710 | 1.57E-65 |
| Peripheral enthesopathies and allied syndromes | 2.35 | 1.89E-02 | 10.22 | 1.54E-24 | 9.32 | 1.13E-20 |
| Reflux esophagitis | 4.41 | 1.05E-05 | 12.30 | 9.58E-35 | 12.08 | 1.31E-33 |
| Tobacco use disorder | 11.52 | 1.06E-30 | 44.62 | 0.00E+00 | 40.53 | 0.00E+00 |
| Transient cerebral ischemia | 7.2975 | 2.93E-13 | 22.62 | 2.68E-113 | 22.46 | 1.10E-111 |
| Type 2 diabetes | 23.13 | 2.25E-118 | 43.41 | 0.00E+00 | 43.12 | 0.00E+00 |
| Ulceration of intestine | 2.25 | 2.46E-02 | 5.95 | 2.63E-09 | 5.3 | 7.02E-08 |
| Urinary tract infection | 2.52 | 1.17E-02 | 16.33 | 5.97E-60 | 13.76 | 4.29E-43 |
| Uveitis, noninfectious or NOS | 10.39 | 2.82E-25 | 14.07 | 5.88E-45 | 14.39 | 6.10E-47 |
| Varicose veins of lower extremity | 8.91 | 5.32E-19 | 10.24 | 1.35E-24 | 10.46 | 1.39E-25 |

Table S13. Difference in PRS rank between emerging cases and persistent controls for the binary, liability and meta methods. This table contains the numerical values of Figure 6A.

| Disease/Trait | PRS Percentile Delta (liability-binary) | PRS Percentile Delta (meta-binary) |
|---|---|---|
| Abdominal pain | 4.81 | 2.73 |
| Alcoholism | 7.05 | 4.65 |
| Allergy/adverse effect of penicillin | 3.47 | 1.71 |
| Angina pectoris | 3.91 | 2.91 |
| Asthma | 4.83 | 4.21 |
| Atrial fibrillation and flutter | 1.06 | 0.54 |
| Benign neoplasm of colon | 1.16 | 0.46 |
| Benign neoplasm of skin | 1.67 | 1.17 |
| Cancer of brain | 1.66 | 0.92 |
| Cataract | 1.34 | 1.01 |
| Cholelithiasis | 1.52 | 1.04 |
| Circulatory disease NEC | 4.27 | 3.09 |
| Coronary atherosclerosis | 1.71 | 1.28 |
| Diaphragmatic hernia | 4.28 | 2.69 |
| Disorders of iris and ciliary body | 2.44 | 1.86 |
| Diverticulosis | 1.51 | 0.91 |
| Enthesopathy | 1.61 | 1.00 |
| Essential hypertension | 3.59 | 3.08 |
| Gastritis and duodenitis | 4.72 | 2.99 |
| Hematuria | 1.50 | 1.26 |
| Hemorrhoids | 1.02 | 0.84 |
| Hypercholesterolemia | 3.12 | 1.72 |
| Hyperplasia of prostate | 8.29 | 6.04 |
| Hypothyroidism NOS | 3.30 | 2.75 |
| Inguinal hernia | 0.86 | 0.36 |
| Malignant neoplasm of female breast | 1.78 | 0.89 |
| Myocardial infarction | 1.69 | 1.70 |
| Obesity | 8.40 | 6.52 |
| Osteoarthritis; localized | 3.17 | 2.29 |
| Peripheral enthesopathies and allied syndromes | 2.85 | 2.23 |
| Reflux esophagitis | 3.81 | 2.30 |
| Tobacco use disorder | 9.01 | 7.01 |
| Transient cerebral ischemia | 3.40 | 2.30 |
| Type 2 diabetes | 4.29 | 3.79 |
| Ulceration of intestine | 1.71 | 1.05 |
| Urinary tract infection | 3.99 | 2.18 |
| Uveitis, noninfectious or NOS | 1.66 | 1.12 |
| Varicose veins of lower extremity | 1.11 | 0.87 |

Table S14. Difference in PRS percentile of emerging cases per trait between liability and binary as well as meta and binary methods. PRS percentiles were calculated relative to the entire dataset and the difference displayed here was calculated on newly diagnosed cases. This table accompanies Figure 6B.

| Disease/Trait | Emerging cases Median Rank (Binary) | Emerging cases Median Rank (Liability) | Emerging cases Median Rank (Meta) |
|---|---|---|---|
| Abdominal pain | 118664 | 135796 | 129592 |
| Alcoholism | 128808 | 157287 | 151198 |
| Allergy/adverse effect of penicillin | 129181 | 140340 | 136639 |
| Angina pectoris | 134134 | 147737 | 144949 |
| Asthma | 133899 | 154813 | 152617 |
| Atrial fibrillation and flutter | 135408 | 141847 | 138161 |
| Benign neoplasm of colon | 130784 | 134240 | 132783 |
| Benign neoplasm of skin | 130659 | 139543 | 137737 |
| Cancer of brain | 127930 | 134467 | 132341 |
| Cataract | 129676 | 135422 | 134286 |
| Cholelithiasis | 133244 | 142432 | 142650 |
| Circulatory disease NEC | 130772 | 147553 | 143118 |
| Coronary atherosclerosis | 135818 | 141436 | 139835 |
| Diaphragmatic hernia | 128108 | 142837 | 137396 |
| Disorders of iris and ciliary body | 124764 | 132882 | 131380 |
| Diverticulosis | 132482 | 136694 | 134371 |
| Enthesopathy | 130572 | 139522 | 135672 |
| Essential hypertension | 114204 | 128652 | 125433 |
| Gastritis and duodenitis | 126143 | 143311 | 138582 |
| Hematuria | 130335 | 133360 | 132446 |
| Hemorrhoids | 124687 | 126685 | 124787 |
| Hypercholesterolemia | 130256 | 142812 | 138410 |
| Hyperplasia of prostate | 123736 | 139824 | 135198 |
| Hypothyroidism NOS | 148651 | 163185 | 161784 |
| Inguinal hernia | 129303 | 133172 | 130400 |
| Malignant neoplasm of female breast | 142973 | 145840 | 144306 |
| Myocardial infarction | 135576 | 144965 | 143320 |
| Obesity | 140675 | 174998 | 169371 |
| Osteoarthritis; localized | 135537 | 145877 | 145849 |
| Peripheral enthesopathies and allied syndromes | 130894 | 140844 | 138958 |
| Reflux esophagitis | 132951 | 144561 | 142355 |
| Tobacco use disorder | 139380 | 175989 | 169326 |
| Transient cerebral ischemia | 109124 | 122170 | 117461 |
| Type 2 diabetes | 142685 | 159876 | 157824 |
| Ulceration of intestine | 129794 | 136510 | 133814 |
| Urinary tract infection | 128677 | 142939 | 139369 |
| Uveitis, noninfectious or NOS | 135431 | 140333 | 139108 |
| Varicose veins of lower extremity | 140980 | 144503 | 143582 |

Table S15. PRS rank of emerging cases in binary, liability and meta methods.

| Disease/Trait | Wilcoxon statistic (liability-binary) | P-value (liability-binary) | Wilcoxon statistic (meta-binary) | P-value (meta-binary) |
|---|---|---|---|---|
| Abdominal pain | 8.20E+07 | 1.51E-119 | 8.59E+07 | 9.00E-77 |
| Alcoholism | 9.96E+06 | 1.01E-66 | 1.02E+07 | 1.45E-58 |
| Allergy/adverse effect of penicillin | 3.17E+07 | 7.65E-26 | 3.27E+07 | 4.62E-15 |
| Angina pectoris | 2.54E+07 | 1.79E-51 | 2.56E+07 | 2.03E-46 |
| Asthma | 3.38E+07 | 7.66E-196 | 3.39E+07 | 6.28E-191 |
| Atrial fibrillation and flutter | 1.11E+07 | 1.48E-03 | 1.13E+07 | 1.08E-01 |
| Benign neoplasm of colon | 9.77E+07 | 2.62E-09 | 1.00E+08 | 3.02E-03 |
| Benign neoplasm of skin | 2.84E+06 | 2.43E-04 | 2.84E+06 | 1.87E-04 |
| Cancer of brain | 1.27E+08 | 9.00E-23 | 1.30E+08 | 5.41E-14 |
| Cataract | 8.59E+07 | 7.88E-15 | 8.66E+07 | 3.91E-12 |
| Cholelithiasis | 7.35E+06 | 2.25E-06 | 7.34E+06 | 1.86E-06 |
| Circulatory disease NEC | 6.48E+07 | 3.83E-86 | 6.65E+07 | 3.83E-65 |
| Coronary atherosclerosis | 4.52E+07 | 3.16E-23 | 4.60E+07 | 7.04E-17 |
| Diaphragmatic hernia | 6.80E+07 | 7.83E-84 | 6.96E+07 | 2.02E-65 |
| Disorders of iris and ciliary body | 6.16E+07 | 1.98E-32 | 6.23E+07 | 1.30E-26 |
| Diverticulosis | 1.56E+08 | 5.34E-15 | 1.58E+08 | 7.45E-09 |
| Enthesopathy | 8.01E+06 | 4.54E-06 | 8.13E+06 | 2.93E-04 |
| Essential hypertension | 3.26E+08 | 0.00E+00 | 3.36E+08 | 0.00E+00 |
| Gastritis and duodenitis | 2.31E+07 | 1.20E-54 | 2.36E+07 | 4.75E-43 |
| Hematuria | 1.29E+07 | 2.17E-06 | 1.30E+07 | 5.97E-06 |
| Hemorrhoids | 1.11E+07 | 2.02E-03 | 1.13E+07 | 6.63E-02 |
| Hypercholesterolemia | 1.33E+08 | 2.72E-91 | 1.39E+08 | 4.48E-54 |
| Hyperplasia of prostate | 1.23E+07 | 4.09E-38 | 1.24E+07 | 2.37E-35 |
| Hypothyroidism NOS | 1.90E+07 | 4.40E-67 | 1.89E+07 | 1.14E-69 |
| Inguinal hernia | 1.92E+07 | 5.58E-03 | 1.94E+07 | 5.80E-02 |
| Malignant neoplasm of female breast | 2.96E+06 | 8.29E-05 | 3.12E+06 | 1.62E-01 |
| Myocardial infarction | 2.30E+07 | 1.65E-24 | 2.31E+07 | 1.78E-22 |
| Obesity | 4.62E+07 | 0.00E+00 | 4.57E+07 | 0.00E+00 |
| Osteoarthritis; localized | 1.81E+07 | 8.75E-20 | 1.80E+07 | 1.23E-20 |
| Peripheral enthesopathies and allied syndromes | 5.11E+06 | 1.80E-12 | 5.11E+06 | 1.76E-12 |
| Reflux esophagitis | 9.10E+06 | 6.95E-17 | 9.15E+06 | 7.40E-16 |
| Tobacco use disorder | 1.32E+07 | 2.43E-202 | 1.33E+07 | 2.07E-193 |
| Transient cerebral ischemia | 2.00E+08 | 4.73E-196 | 2.10E+08 | 8.80E-125 |
| Type 2 diabetes | 3.88E+07 | 2.14E-112 | 3.88E+07 | 1.28E-111 |
| Ulceration of intestine | 5.31E+06 | 7.06E-05 | 5.40E+06 | 1.95E-03 |
| Urinary tract infection | 3.23E+07 | 1.31E-41 | 3.32E+07 | 5.02E-29 |
| Uveitis, noninfectious or NOS | 4.50E+07 | 1.32E-11 | 4.51E+07 | 8.98E-11 |
| Varicose veins of lower extremity | 3.10E+06 | 2.79E-03 | 3.13E+06 | 9.96E-03 |

Table S16. Difference in PRS rank of emerging cases between liability and binary methods as well as between meta and binary methods.

| Disease/Trait | Wilcoxon statistic (Binary) | P-value (Binary) | Wilcoxon statistic (Liability) | P-value (Liability) | Wilcoxon statistic (Meta) | P-value (Meta) |
|---|---|---|---|---|---|---|
| Abdominal pain | 3.58 | 3.48E-04 | 18.44 | 6.23E-76 | 16.7 | 1.40E-62 |
| Alcoholism | 1.85 | 6.48E-02 | 22.51 | 3.44E-112 | 19.26 | 1.20E-82 |
| Allergy/adverse effect of penicillin | 4.43 | 9.64E-06 | 12.53 | 4.88E-36 | 11.79 | 4.18E-32 |
| Angina pectoris | 9.82 | 9.56E-23 | 22.6 | 4.22E-113 | 21.46 | 3.57E-102 |
| Asthma | 14.9 | 3.35E-50 | 37.31 | 1.29E-304 | 36.18 | 1.44E-286 |
| Atrial fibrillation and flutter | 8.16 | 3.24E-16 | 10.72 | 7.96E-27 | 10.24 | 1.27E-24 |
| Benign neoplasm of colon | 12.26 | 1.46E-34 | 14.23 | 6.18E-46 | 13.84 | 1.42E-43 |
| Benign neoplasm of skin | 1.76 | 7.79E-02 | 5.49 | 3.96E-08 | 5.42 | 5.95E-08 |
| Cancer of brain | 7.71 | 1.28E-14 | 14.01 | 1.40E-44 | 13.41 | 5.59E-41 |
| Cataract | 6.76 | 1.36E-11 | 12.89 | 4.98E-38 | 13.06 | 5.91E-39 |
| Cholelithiasis | 5 | 5.63E-07 | 10.34 | 4.50E-25 | 10.64 | 1.95E-26 |
| Circulatory disease NEC | 7.48 | 7.35E-14 | 28.19 | 6.95E-175 | 25.12 | 3.31E-139 |
| Coronary atherosclerosis | 14.19 | 1.12E-45 | 20.36 | 3.51E-92 | 19.81 | 2.47E-87 |
| Diaphragmatic hernia | 11.11 | 1.18E-28 | 24.74 | 3.63E-135 | 24.07 | 5.57E-128 |
| Disorders of iris and ciliary body | 4.95 | 7.31E-07 | 12.63 | 1.41E-36 | 12.59 | 2.36E-36 |
| Diverticulosis | 21.35 | 3.76E-101 | 22.24 | 1.30E-109 | 24.09 | 3.28E-128 |
| Enthesopathy | 3.6 | 3.23E-04 | 7.93 | 2.19E-15 | 7.08 | 1.47E-12 |
| Essential hypertension | 23.21 | 3.68E-119 | 45.18 | 0.00E+00 | 45.04 | 0.00E+00 |
| Gastritis and duodenitis | 1.64 | 1.02E-01 | 15.76 | 6.23E-56 | 13.76 | 4.64E-43 |
| Hematuria | 3.46 | 5.41E-04 | 6.9 | 5.09E-12 | 6.86 | 6.78E-12 |
| Hemorrhoids | 2.54 | 1.12E-02 | 2.35 | 1.85E-02 | 2.76 | 5.73E-03 |
| Hypercholesterolemia | 17.74 | 1.91E-70 | 33.62 | 7.76E-248 | 31.35 | 9.62E-216 |
| Hyperplasia of prostate | 6.87 | 6.33E-12 | 14.17 | 1.37E-45 | 13.95 | 2.99E-44 |
| Hypothyroidism NOS | 21.91 | 2.31E-106 | 35.04 | 5.65E-269 | 33.98 | 4.91E-253 |
| Inguinal hernia | 5.81 | 6.25E-09 | 6 | 2.02E-09 | 6.52 | 6.86E-11 |
| Malignant neoplasm of female breast | 12.07 | 1.45E-33 | 14.06 | 6.24E-45 | 12.84 | 1.02E-37 |
| Myocardial infarction | 10.51 | 7.50E-26 | 17.78 | 1.05E-70 | 17.99 | 2.50E-72 |
| Obesity | 18.66 | 1.04E-77 | 60.54 | 0.00E+00 | 55.13 | 0.00E+00 |
| Osteoarthritis; localized | 7.79 | 6.94E-15 | 17.2 | 2.61E-66 | 16.61 | 6.08E-62 |
| Peripheral enthesopathies and allied syndromes | 2.35 | 1.89E-02 | 8.89 | 6.35E-19 | 8.44 | 3.28E-17 |
| Reflux esophagitis | 4.41 | 1.05E-05 | 12.52 | 5.94E-36 | 11.98 | 4.78E-33 |
| Tobacco use disorder | 11.52 | 1.06E-30 | 42.9 | 0.00E+00 | 38.57 | 0.00E+00 |
| Transient cerebral ischemia | 7.3 | 2.93E-13 | 16.69 | 1.58E-62 | 17.01 | 6.64E-65 |
| Type 2 diabetes | 23.13 | 2.25E-118 | 41.52 | 0.00E+00 | 41.37 | 0.00E+00 |
| Ulceration of intestine | 2.25 | 2.46E-02 | 5.34 | 9.19E-08 | 4.69 | 2.79E-06 |
| Urinary tract infection | 2.52 | 1.17E-02 | 16.43 | 1.11E-60 | 13.97 | 2.23E-44 |
| Uveitis, noninfectious or NOS | 10.39 | 2.82E-25 | 14.89 | 3.90E-50 | 15.42 | 1.16E-53 |
| Varicose veins of lower extremity | 8.91 | 5.32E-19 | 10.41 | 2.30E-25 | 10.24 | 1.28E-24 |
| Urinary tract infection | 2.11 | 3.52E-02 | 13.25 | 4.30E-40 | 11.75 | 6.71E-32 |
| Uveitis, noninfectious or NOS | 8.23 | 1.85E-16 | 13.53 | 1.03E-41 | 14.47 | 2.01E-47 |
| Varicose veins of lower extremity | 7.45 | 9.07E-14 | 8.33 | 7.56E-17 | 8.48 | 2.33E-17 |

Table S17. Difference in PRS between emerging cases and persistent controls for the binary, liability and meta methods calculated in an independent test set. This table contains the numerical values of Figure S6 and S7.

| Disease/Trait | Emerging cases Median Rank (Binary) | Emerging cases Median Rank (Liability) | Emerging cases Median Rank (Meta) |
|---|---|---|---|
| Abdominal pain | 24271 | 27760 | 26266 |
| Alcoholism | 25959 | 30095 | 29194 |
| Allergy/adverse effect of penicillin | 25674 | 28440 | 27486 |
| Angina pectoris | 25443 | 29180 | 28826 |
| Asthma | 26637 | 30613 | 30408 |
| Atrial fibrillation and flutter | 26264 | 28595 | 27699 |
| Benign neoplasm of colon | 25973 | 27504 | 26971 |
| Benign neoplasm of skin | 26389 | 28532 | 28101 |
| Cancer of brain | 25632 | 26871 | 26365 |
| Cataract | 25774 | 27065 | 26377 |
| Cholelithiasis | 26385 | 27426 | 27171 |
| Circulatory disease NEC | 26070 | 29299 | 28409 |
| Coronary atherosclerosis | 27075 | 28315 | 28575 |
| Diaphragmatic hernia | 26310 | 28499 | 27836 |
| Disorders of iris and ciliary body | 24732 | 27010 | 26263 |
| Diverticulosis | 26311 | 27232 | 26912 |
| Enthesopathy | 25399 | 26944 | 27491 |
| Essential hypertension | 23761 | 26172 | 25890 |
| Gastritis and duodenitis | 26381 | 28678 | 27595 |
| Hematuria | 24832 | 27517 | 26695 |
| Hemorrhoids | 25652 | 26345 | 25716 |
| Hypercholesterolemia | 26259 | 28761 | 28229 |
| Hyperplasia of prostate | 12121 | 12460 | 12598 |
| Hypothyroidism NOS | 30003 | 32425 | 32279 |
| Inguinal hernia | 26376 | 26439 | 26143 |
| Malignant neoplasm of female breast | 15945 | 15899 | 15503 |
| Myocardial infarction | 27598 | 29393 | 28917 |
| Obesity | 28232 | 33484 | 32774 |
| Osteoarthritis; localized | 26709 | 29129 | 28904 |
| Peripheral enthesopathies and allied syndromes | 26177 | 27944 | 27827 |
| Reflux esophagitis | 26428 | 27925 | 28678 |
| Tobacco use disorder | 28278 | 33676 | 33020 |
| Transient cerebral ischemia | 23017 | 25387 | 24325 |
| Type 2 diabetes | 28353 | 31570 | 31258 |
| Ulceration of intestine | 27060 | 28771 | 28384 |
| Urinary tract infection | 26373 | 28329 | 27579 |
| Uveitis, noninfectious or NOS | 27754 | 28010 | 28096 |
| Varicose veins of lower extremity | 27998 | 27215 | 26954 |

Table S18. PRS rank of emerging cases in binary, liability and meta methods calculated in an independent test set.

| Disease/Trait | Wilcoxon statistic (liability-binary) | P-value (liability-binary) | Wilcoxon statistic (meta-binary) | P-value (meta-binary) |
|---|---|---|---|---|
| Abdominal pain | 3.53E+06 | 4.28E-19 | 3.64E+06 | 6.11E-14 |
| Alcoholism | 4.36E+05 | 7.49E-07 | 4.36E+05 | 6.00E-07 |
| Allergy/adverse effect of penicillin | 1.30E+06 | 4.16E-08 | 1.35E+06 | 2.57E-05 |
| Angina pectoris | 9.76E+05 | 2.31E-15 | 9.87E+05 | 3.90E-14 |
| Asthma | 1.49E+06 | 1.34E-30 | 1.51E+06 | 2.12E-28 |
| Atrial fibrillation and flutter | 4.08E+05 | 9.13E-05 | 4.05E+05 | 3.73E-05 |
| Benign neoplasm of colon | 3.87E+06 | 7.65E-03 | 3.96E+06 | 1.20E-01 |
| Benign neoplasm of skin | 1.22E+05 | 2.08E-01 | 1.21E+05 | 1.44E-01 |
| Cancer of brain | 4.98E+06 | 2.07E-05 | 5.05E+06 | 4.64E-04 |
| Cataract | 3.55E+06 | 1.31E-02 | 3.60E+06 | 7.80E-02 |
| Cholelithiasis | 3.15E+05 | 1.69E-01 | 3.18E+05 | 2.67E-01 |
| Circulatory disease NEC | 2.72E+06 | 3.92E-13 | 2.78E+06 | 3.52E-10 |
| Coronary atherosclerosis | 1.89E+06 | 5.10E-05 | 1.90E+06 | 9.17E-05 |
| Diaphragmatic hernia | 2.91E+06 | 1.68E-11 | 2.95E+06 | 9.72E-10 |
| Disorders of iris and ciliary body | 2.42E+06 | 1.78E-06 | 2.45E+06 | 2.26E-05 |
| Diverticulosis | 6.33E+06 | 4.18E-04 | 6.39E+06 | 3.21E-03 |
| Enthesopathy | 3.51E+05 | 8.34E-02 | 3.42E+05 | 1.51E-02 |
| Essential hypertension | 1.39E+07 | 2.76E-57 | 1.42E+07 | 2.83E-46 |
| Gastritis and duodenitis | 9.56E+05 | 5.45E-05 | 9.61E+05 | 1.07E-04 |
| Hematuria | 5.43E+05 | 8.80E-04 | 5.52E+05 | 4.89E-03 |
| Hemorrhoids | 4.62E+05 | 1.96E-01 | 4.71E+05 | 5.05E-01 |
| Hypercholesterolemia | 5.80E+06 | 3.72E-13 | 5.99E+06 | 5.26E-08 |
| Hyperplasia of prostate | 5.55E+05 | 4.50E-03 | 5.37E+05 | 1.26E-04 |
| Hypothyroidism NOS | 8.01E+05 | 3.58E-08 | 7.79E+05 | 1.41E-10 |
| Inguinal hernia | 7.46E+05 | 7.75E-01 | 7.30E+05 | 2.91E-01 |
| Malignant neoplasm of female breast | 1.42E+05 | 5.23E-01 | 1.42E+05 | 5.53E-01 |
| Myocardial infarction | 9.97E+05 | 7.47E-03 | 9.93E+05 | 4.67E-03 |
| Obesity | 2.01E+06 | 8.75E-43 | 1.99E+06 | 1.42E-45 |
| Osteoarthritis; localized | 7.35E+05 | 5.77E-04 | 7.27E+05 | 1.56E-04 |
| Peripheral enthesopathies and allied syndromes | 1.93E+05 | 1.27E-01 | 1.93E+05 | 1.10E-01 |
| Reflux esophagitis | 3.74E+05 | 1.42E-03 | 3.72E+05 | 9.67E-04 |
| Tobacco use disorder | 6.43E+05 | 2.48E-25 | 6.42E+05 | 1.40E-25 |
| Transient cerebral ischemia | 8.48E+06 | 9.40E-23 | 8.82E+06 | 1.41E-13 |
| Type 2 diabetes | 1.64E+06 | 2.23E-17 | 1.61E+06 | 7.58E-20 |
| Ulceration of intestine | 2.04E+05 | 2.06E-01 | 2.02E+05 | 1.25E-01 |
| Urinary tract infection | 1.44E+06 | 1.15E-04 | 1.45E+06 | 4.43E-04 |
| Uveitis, noninfectious or NOS | 1.84E+06 | 9.38E-02 | 1.81E+06 | 1.88E-02 |
| Varicose veins of lower extremity | 1.34E+05 | 8.75E-01 | 1.34E+05 | 7.46E-01 |

Table S19. Difference in PRS rank of emerging cases between liability and binary methods as well as between meta and binary methods calculated in an independent test set.
